# Leukocytes carrying *Clonal Hematopoiesis of Indeterminate Potential* (CHIP) Mutations invade Human Atherosclerotic Plaques

**DOI:** 10.1101/2023.07.22.23292754

**Authors:** Moritz von Scheidt, Sabine Bauer, Angela Ma, Ke Hao, Thorsten Kessler, Baiba Vilne, Ying Wang, Chani J. Hodonsky, Saikat K.B. Ghosh, Michal Mokry, Hua Gao, Kenji Kawai, Atsushi Sakamoto, Juliane Kaiser, Dario Bongiovanni, Julia Fleig, Lilith Oldenbuettel, Zhifen Chen, Aldo Moggio, Hendrik B. Sager, Judith S. Hecker, Florian Bassermann, Lars Maegdefessel, Clint L. Miller, Wolfgang Koenig, Andreas M. Zeiher, Stefanie Dimmeler, Matthias Graw, Christian Braun, Arno Ruusalepp, Nicholas J. Leeper, Jason C. Kovacic, Johan L.M. Björkegren, Heribert Schunkert

**Author notes:** Corresponding authors: Prof. Dr. med. Heribert Schunkert, MD; and Dr. Dr. med. Moritz von Scheidt, MD; Deutsches Herzzentrum München (German Heart Center Munich); Klinik für Herz- und Kreislauferkrankungen Lazarettstr. 36; D-80636 München. Authors contributed equally (Johan L.M. Björkegren, Heribert Schunkert).

## Abstract

**Background:** Leukocyte progenitors derived from clonal hematopoiesis of undetermined potential (CHIP) are associated with increased cardiovascular events. However, the prevalence and functional relevance of CHIP in coronary artery disease (CAD) are unclear, and cells affected by CHIP have not been detected in human atherosclerotic plaques.

**Methods:** CHIP mutations in blood and tissues were identified by targeted deep-DNA-sequencing (DNAseq: coverage >3,000) and whole-genome-sequencing (WGS: coverage >35). CHIP-mutated leukocytes were visualized in human atherosclerotic plaques by mutaFISH^TM^. Functional relevance of CHIP mutations was studied by RNAseq.

**Results:** DNAseq of whole blood from 540 deceased CAD patients of the Munich cardIovaScular StudIes biObaNk (MISSION) identified 253 (46.9%) CHIP mutation carriers (mean age 78.3 years). DNAseq on myocardium, atherosclerotic coronary and carotid arteries detected identical CHIP mutations in 18 out of 25 mutation carriers in tissue DNA. MutaFISH^TM^ visualized individual macrophages carrying *DNMT3A* CHIP mutations in human atherosclerotic plaques. Studying monocyte-derived macrophages from Stockholm-Tartu Atherosclerosis Reverse Networks Engineering Task (STARNET; n=941) by WGS revealed CHIP mutations in 14.2% (mean age 67.1 years). RNAseq of these macrophages revealed that expression patterns in CHIP mutation carriers differed substantially from those of non-carriers. Moreover, patterns were different depending on the underlying mutations, e.g. those carrying *TET2* mutations predominantly displayed upregulated inflammatory signaling whereas *ASXL1* mutations showed stronger effects on metabolic pathways.

**Conclusions:** Deep-DNA-sequencing reveals a high prevalence of CHIP mutations in whole blood of CAD patients. CHIP-affected leukocytes invade plaques in human coronary arteries. RNAseq data obtained from macrophages of CHIP-affected patients suggest that pro-atherosclerotic signaling differs depending on the underlying mutations. Further studies are necessary to understand whether specific pathways affected by CHIP mutations may be targeted for personalized treatment.

## Introduction

Clonal hematopoiesis of undetermined potential (CHIP) is a common age-related condition and potential precursor to hematological neoplasms. CHIP is defined as presence of a clonally expanded hematopoietic cell caused by a somatic mutation in individuals without hematologic abnormalities. Such mutations, predominantly in the epigenetic regulators *DNMT3A*, *TET2* or *ASXL1*, have been identified in bone marrow and blood, but these cells have never been definitively detected in human atherosclerotic plaques.^1^ Whole-exome sequencing has shown that CHIP is practically absent in individuals younger than 30 years of age, whereas it is present in 20% to 30% of individuals aged 50 to 60 years.^1, 2^ The use of more sensitive sequencing techniques and access to bone marrow has demonstrated that small clones are quite common in middle-aged individuals.^3, 4^ CHIP carriers have an increased risk of cardiovascular events and mortality, including early onset and aggravated progression of coronary artery disease (CAD), myocardial infarction, stroke, aortic valve calcification and chronic ischemic heart failure.^5–8^ The higher the burden of CHIP-affected cells measured by the variant allele frequency (VAF), the higher was the reported risk of adverse events.^5–8^ Indeed, detection of CHIP in peripheral blood may serve as a biomarker for adverse outcomes in individuals with cardiovascular diseases (CVDs).

Experimental studies deciphered the first mechanistic insights explaining the associations of CHIP mutations with CVD. Fuster et al. demonstrated that transplantation of bone marrow containing a *Tet2* loss-of-function mutation in hematopoietic cells in mice mimics the human situation of CHIP including increased numbers of activated macrophages (measured by upregulation of *IL-1β*) found in plaques. Aggravated atherosclerosis was triggered by activation of the inflammatory cascade in these cells. Further, *in-vitro* LPS-stimulation of macrophages carrying somatic mutations led to increased expression of *IL-6*, the downstream mediator of the inflammasome complex, and *IL-1β*.^9^

Our current understanding is that driver mutations in *DNMT3A*, *TET2* or *ASXL1* lead to expansion of clones in the bone marrow resulting in increased numbers of cells with epigenetic alterations in the peripheral blood. These changes result in increased expression of genes associated with inflammatory pathways, which in turn may stimulate the progression of atherosclerosis.^10^ Here we studied the prevalence of CHIP in DNA sequence data, whether CHIP-mutated leukocytes invade atherosclerotic plaques in human arteries and whether macrophages isolated from CAD patients carrying specific CHIP mutations also present with specific CAD signatures by examining macrophage RNA-expression profiles and associated clinical phenotypes (**Figure 1**).

**Figure 1.**
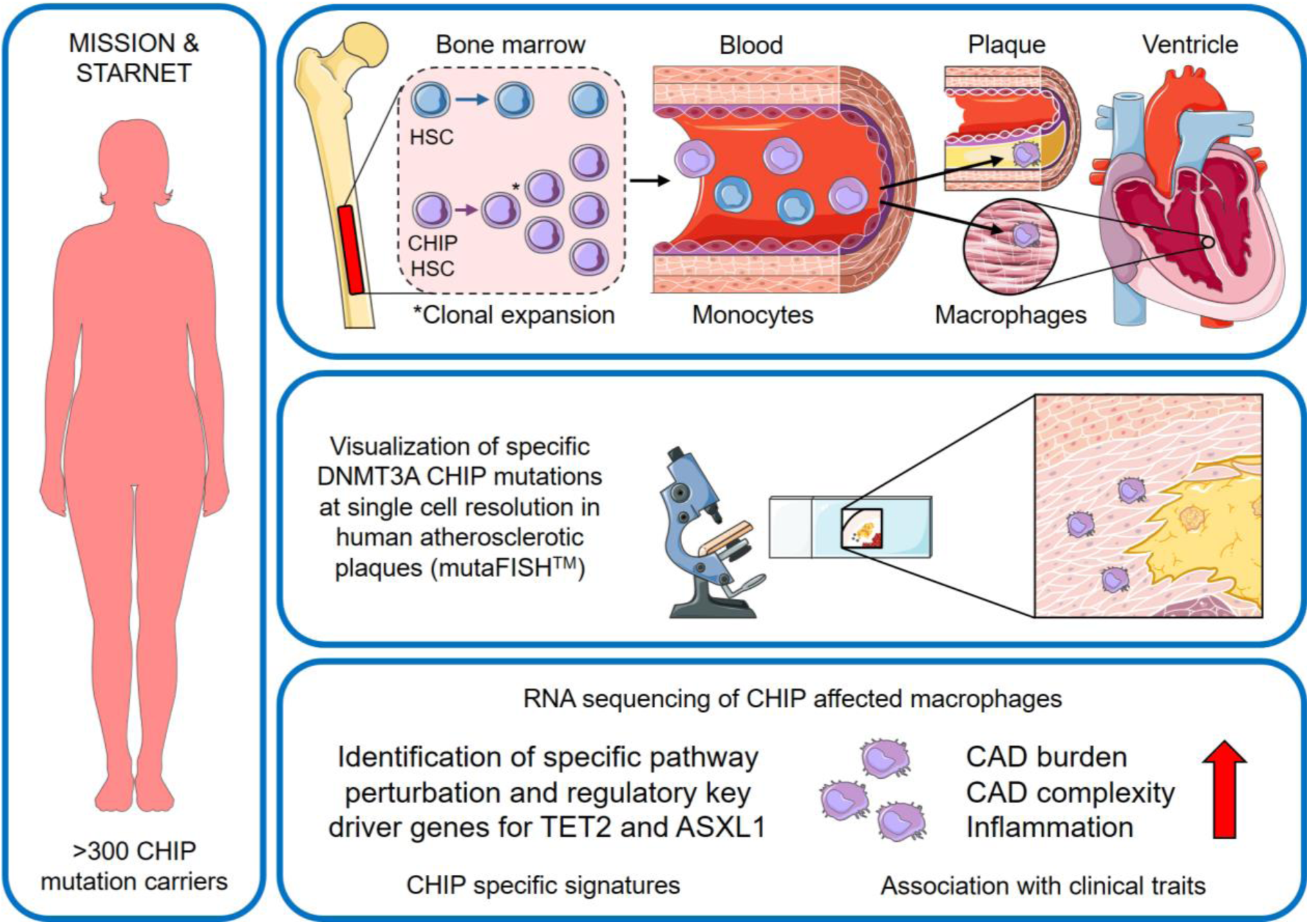
Central illustration – important novel findings from this study. **Left panel:** Over 300 CHIP mutation carriers from the Munich cardIovaScular StudIes biObaNk (MISSION) and the Stockholm-Tartu Atherosclerosis Reverse Networks Engineering Task (STARNET) studies were evaluated. **Upper panel**: In our study, we confirmed blood derived CHIP mutations in human atherosclerotic plaques of coronary artery, carotids and heart muscle on DNA level. **Middle panel:** For the first time we were able to visualize macrophages with specific *DNMT3A* CHIP mutations using mutaFISH^TM^ at single cell resolution in human atherosclerotic plaques. Enrichment of CHIP mutated cells in human plaques was confirmed. **Lower panel:** Previously unknown pro-atherosclerotic alterations in gene expression of CHIP mutated macrophages at the level of regulatory key-drivers, pathways, networks, and modules with relevance for CAD progression were identified. ASXL1: ASXL Transcriptional Regulator 1; CAD: Coronar artery disease; CHIP: Clonal hematopoiesis of indeterminate potential; DNMT3A: DNA Methyltransferase 3 Alpha; HSC: hematopoietic stem cell; mutaFISH: mutation-specific Fluorescence In Situ Hybridization; TET2: Tet Methylcytosine Dioxygenase 2.

## Materials and Methods

### Ethics approval

The institutional review board and Ethics Committee of the Technical University of Munich, Germany, approved the protocol of MISSION (2018-325-S-KK – 08/22/2018). The use of human STARNET samples has been approved by the Estonian Bioethics and Human Research Committee (Ministry of Social Affairs) (IRB 2771T,17 – 12/01/2018) and by the written informed consent of the donor, in accordance with the guidelines and regulations for the use of biological material of human origin. Both studies were conducted in accordance with the provisions of the Declaration of Helsinki and the International Conference on Harmonization guidelines for good clinical practice.

### Data accessibility

Data used in this study are available in permanent repositories. Human data from MISSION can be requested by qualified researchers at the German Heart Center Munich from the corresponding author. Human data from STARNET are accessible through the Database of Genotypes and Phenotypes (dbGAP).

### MISSION – cohort and sample description

The Munich cardIovaScular StudIes biObaNk (MISSION) was started in 2019 and comprises cardiovascular relevant tissues, including blood and plasma, liver, myocardium, coronary, and carotid samples from >950 deceased individuals sampled in FFPE and fresh frozen at −80°C. Prior to freezing and sequencing, all tissues are washed and subsequently conserved in PBS/DMSO. MISSION provides the full spectrum of coronary phenotypes from healthy to severe atherosclerosis. DNA of leukocytes and cardiovascular tissues was analyzed by deep-targeted-amplicon-sequencing. The leukocytes derived from whole blood, and the analyzed tissue encompassed the proximal part of atherosclerosis-affected left anterior descending (LAD) coronary, atherosclerosis-affected left carotid artery and left ventricular heart muscle.

### DNA extraction from blood and tissue

For the extraction of DNA from whole blood, the Maxwell DNA Blood Kit (Promega) was used. For this purpose, whole blood was incubated for 20 minutes at room temperature under rotation (Rotating shaker, Kisker Biotech, Germany). Then, 300µl of whole blood was mixed with 300µl of lysis buffer and 30µl of proteinase K. The mixture was incubated under rotation for 30 minutes at room temperature, followed by another 30 minutes at 65°C and 600rpm in a heat block (Thermomixer comfort, Eppendorf, Germany). Every 10 minutes, incubation was performed at 1500rpm for approximately 1 minute. DNA extraction from human coronary artery, carotid and left ventricle was performed based on 50mg of frozen tissue samples. Perivascular tissue was removed from arteries and tissues were washed in 1x PBS. Isolation was performed based on a modified Maxwell DNA Blood kit (Promega) protocol. After mechanical homogenization (TissueLyser II, Qiagen, The Netherlands) of the tissues in 300µl incubation buffer, 1-thioglycerol and proteinase K were added. The mixture was incubated for 4 hours at 65°C and 600rpm in a heat block (Thermomixer comfort, Eppendorf, Germany). This was followed by the addition of 300µl of lysis buffer and re-incubation for 10 minutes at 600rpm. Finally, the batches (blood and tissue) were transferred to the first well of the cartridge included in the kit. After addition of the plunger, the cartridge was inserted into the Maxwell RSC 48 system. The supplied elution tubes (0.5ml) were filled with 65µl of elution buffer and also inserted into the instrument. Finally, the predefined Maxwell DNA Blood protocol was selected and DNA was isolated in around 37 minutes. Isolated DNA was measured fluorometrically with a Qubit 3.0 (ThermoScientific).

### Sample preparation and DNA extraction

Whole blood and cardiovascular tissues were analyzed by deep-DNA-sequencing to detect hematopoietic stem cell derived mononuclear cells with CHIP mutations in cardiovascular relevant tissues. Samples were sequenced using an Illumina TruSeq Custom sequencing panel at the Munich Leukemia Laboratory. The concentration of extracted genomic DNA was determined with a Qubit dsDNA HS assay kit (Life Technologies) and then diluted to 25ng/µl in 30µl of nuclease free water. A depth of more than 1000 detections per gene was targeted for sequencing to reliably detect CHIP-mutated hematopoietic cells in whole blood, atherosclerosis-affected coronary and carotid artery, and myocardium of the left ventricle.

### Deep-DNA-sequencing

A previously established panel containing 594 amplicons in 56 genes was used in the Illumina TruSeq Custom Amplicon Low Input assay.^6^ To allow improved identification of low allele frequency variants, double-strand sequencing was performed. In addition, 6-bp unique molecular identifiers (UMIs) were included in the target-specific primers. Prior to sequencing, pooled libraries were diluted and denatured according to the NextSeq System Denature and Dilute Libraries Guide (Illumina). 1% PhiX DNA was added. Pooled libraries were sequenced with the NextSeq 500 sequencer (Illumina) using the NextSeq 500/550 Mid Output, Version 2 kit (300 cycles) according to the manufacturer’s instructions. The sequencer was run in paired-end sequencing mode with a read length of 2×150bp and an index read length of 2×8bp. The BCL files were demultiplexed and converted to FASTQ files using the FASTQ Generation tool on BaseSpace (Illumina). The average coverage of the samples was >3,000 per gene. The variants were further validated on the basis of being reported in the literature and/or the Catalogue of Somatic Mutations in Cancer (https://cancer.sanger.ac.uk/cosmic) and ClinVar (https://www.ncbi.nlm.nih.gov/clinvar). The 56 gene panel was adapted after screening the first n=192 individuals to a custom 13 gene panel comprising *ASXL1*, *CALR*, *CBL*, *DNMT3A*, *JAK2*, *MPL*, *PPM1D*, *SF3B1*, *SRSF2*, *TET2*, *TP53*, *U2AF1* and *ZRSR2*, thereby covering almost 90% of known CHIP mutations. The 13 gene panel followed the same analysis pipeline.

### Visualization of specific CHIP mutations in human plaques

Mutation specific Fluorescence In Situ Hybridization (mutaFISH^TM^) analysis on *DNMT3A* gene was performed on 3-5µm thick tissue sections from human coronary and carotid arteries of patients with known mutation (c.2245C>T and c.2333T>G) in the *DNMT3A* gene. In situ rolling circle technology was performed using custom made dual color *DNMT3A* mutation and wildtype specific FISH probes (Abnova, Taiwan). The detailed protocol used for pretreatment, reverse transcription, hybridization, amplification and signal detection is provided in the supplement. It is based on the manufacturers protocol for mutaFISH^TM^ RNA Accessory Kit and the protocol for muta FISH^TM^ HER2wt RNA probes with some modifications in target retrieval, washing buffer and incubation. Negative controls were used to establish unspecific binding and background. All reagents were prepared with RNAse free (DEPC treated) water/PBS. For hybridization a Boekel Scientific Slide Moat oven was used. Images were acquired at the ZEISS Axioscan 7 slide scanner and analysis was performed using Zeiss Software ZEN 3.5 blue edition.

### STARNET – Study and sample description

To further study the ability of CHIP-mutated leukocytes to invade atherosclerotic plaques and to identify their biological impact we leveraged the Stockholm-Tartu Atherosclerosis Reverse Networks Engineering Task (STARNET) study datasets. Briefly, patients with CAD undergoing coronary artery bypass grafting (CABG; n=941) donated multiple tissue samples that included liver, skeletal muscle, subcutaneous fat, visceral abdominal fat, atherosclerotic aortic wall, internal mammary artery and whole blood as previously described.^11^ After quality control, WGS data of all 941 STARNET individuals were suitable for downstream CHIP analysis.

### Whole genome sequencing

DNA from whole blood was isolated with the QIAmp DNA Blood Midi kit (Qiagen). DNA qualities were assessed with the Agilent 2100 Bioanalyzer system (Agilent Technologies, Palo Alto, CA). Library preparation and sequencing were performed at Beijing Genomic Institute (BGI). Genomic DNA samples that passed quality control were randomly fragmented by Covaris technology and 350bp fragments were selected. End repair of DNA fragments was performed and an “A” base was added at the 3’-end of each strand. Adapters were then ligated to both ends of the end repaired/dA tailed DNA fragments and amplified by ligation-mediated PCR (LM-PCR), followed by single strand separation and cyclization. Rolling circle amplification was performed to produce DNA Nanoballs (DNBs). The qualified DNBs were loaded into patterned nanoarrays and pair-end reads were read through on the BGISEQ-500 platform. High-throughput sequencing was performed for each library to ensure that each sample met the average sequencing coverage requirement of 35x. Sequencing-derived raw image files were processed by BGISEQ-500 base-calling software with default parameters, and the sequence data of each individual was generated as paired-end reads.

Raw data in FASTQ format was filtered and raw reads with low quality were removed. Data was aligned to the human reference genome (GRCh37/HG19) by Burrows-Wheeler Aligner (BWA) v0.7.12 ^12, 13^ and variant calling was performed by Genome Analysis Toolkit (v3.3.0)^14^ with duplicate reads removed by Picard tools v1.118.^15^

The HaplotypeCaller of GATK was used to call both SNPs and InDels simultaneously via local de-novo assembly of haplotypes in a region showing signs of variation.^16, 17^ Base quality scores were recalibrated using GATK BaseRecalibrator and SNPs recalibration was performed using GATK VariantRecalibrator function.^18, 19^

### CHIP somatic variant identification and validation

Heterozygous missense, nonsense, InDel and splicing variants of one or two base pairs in coding regions of CHIP-associated genes including *ASXL1, TET2, JAK2,* and *DNMT3A* were identified by filtering data from vcf files according to: 1) genotype quality > 30; 2) minor allele frequency <1% in 1000 genome database; 3) minor allele frequency <1% in the cohort.

Variant analysis in RNAseq data in blood, aorta, liver, skeletal muscle, subcutaneous fat, visceral abdominal fat, monocyte derived macrophages and macrophage derived foam cells from STARNET subjects was performed for identification and validation of somatic mutations identified via whole genome sequencing. Because bone marrow or hematopoietic cells that carry somatic mutations could present in leukocytes in whole blood, in atherosclerotic plaques in the diseased aorta, or in macrophages and foam cells isolated and differentiated from whole blood, variants that presented in at least one of these four tissues/cell types but not in any other tissues (liver, skeletal muscle, subcutaneous fat, visceral abdominal fat) were defined as somatic mutations.

### RNA sequencing and data processing

The effects of CHIP mutations on gene expression were studied in macrophages and foam cells. After centrifugation of whole blood, the cell pellet was washed with 1x PBS and cells were plated. After 3-6 hours, the monocytes adhered to the plate and non-adherent erythrocytes and lymphocytes could be washed off during adhesion purification. The monocytes were subsequently cultured in human serum, stimulated and differentiated into macrophages over 48-72 hours. The transition to foam cells was achieved by treatment with oxidized low-density lipoprotein (LDL) cholesterol. Finally, the macrophages and foam cells were harvested and RNA was isolated.^11^

RNA library preparation was based on the Ribo-Zero library preparation method using Illumina TruSeq nonstranded mRNA kit. Samples were randomized to prevent batch effects. Sequencing was performed with paired-end reads of 100 base pairs on an Illumina HiSeq, and quality control was performed using FASTQC3. GENCODE was used to quantify the expression of genes and isoforms and mapped to the human genome using STAR4. The average coverage was >40 million reads per sample. Only samples with more than 1 million unambiguously assigned reads were used for further analysis.

### Genetics, imputation and eQTL analysis

Genetic data were analyzed on a genome build 37 background using GenomeStudio (Illumina). After genetic sex confirmation, quality control was performed using PLINK v 2.05. Data were imputed using the HRC r1.1 2016 reference panel using minimac4. Cis- and trans-regulated expression quantitative trait loci (eQTLs) in macrophages were determined using R package matrix eQTL v.2.1.1. Adjustments were made for age, sex, BMI and the first five genetic principal components. Cis-regulatory SNPs within 1 megabase of the respective gene were determined by a linear regression model (hg19 genomes). An FDR <5% was considered statistically significant.

### Differential gene expression and pathway analysis

Differences in gene expression between CHIP mutation carriers and controls were investigated using the R package limma. Covariates in the linear regression model were age, sex and BMI. Adjusted p-values <0.05 were considered statistically significant. Differentially expressed genes were analyzed for overrepresentation of genes from genome-wide association studies (GWAS) to CAD using PhenoScanner. Differentially expressed genes were analyzed by binomial test for enrichment of Gene Ontology (GO) terms and by Gene Set Enrichment Analysis (GSEA). The maximum size of a gene set in GSEA was set to 3,000. Bonferroni-corrected p-values <0.05 were considered statistically significant.

### Network and Key Driver Analysis

Regulatory networks were reconstructed using GENIE3, based on a random forests ensemble method.^20^ The network was enriched with transcription factors and cis-eQTL-regulated genes from macrophages as candidate regulators.^21, 22^ Weighted key driver analysis (wKDA) was performed using mergeomics.^23, 24^ Mergeomics allows disease-relevant processes to be mapped onto molecular interaction networks in order to identify hubs as potential key regulators. The network visualization was realized with Cytoscape v3.7.0.

### Co-expression modules and association with clinical traits

Correlation patterns between gene expression were analyzed using R package Weighted Gene Co-expression Network Analysis (WGCNA) to construct correlation networks and identify co-expression modules. Correlations of gene expression with clinical traits were calculated using Spearman correlation. The association of co-expression modules with clinical traits was analyzed using Fisher’s exact test to calculate the enrichment of the number of significantly correlated genes in each module.

## Results

### CHIP screening by deep DNA sequencing

In MISSION, a total of 540 individuals who died at an age between 40 to 98 years old (mean age 75.1 years) were found to have CAD at autopsy. These subjects were screened for CHIP mutations with a mean sequencing depth of 3,592-fold, with at least 1,000-fold per gene. A total of 253 individuals (46.9%) carried 445 individual CHIP mutations in whole blood. The prevalence of CHIP mutations increased with age (R=0.76; p<0.001), the mean age of mutation carriers was 78.3 years and 113 (44.7%) were female. 78 individuals carried at least one CHIP mutation with a VAF >10%, 130 individuals carried at least one CHIP mutation with a VAF >2% and 45 individuals were identified with clonal hematopoiesis with a VAF <2% (**Supplemental Figure 1**). The greatest burden of CHIP was found in an 85-year-old man who carried 6 unique mutations. Most somatic mutations were identified in *DNMT3A* (155 mutations), *TET2* (152) and *ASXL1* (32). Further, CHIP mutations were detected in *BCOR*, *CBL*, *CALR, CBL*, *EZH2*, *GNAS*, *GNB1*, *IDH1*, *JAK2*, *KRAS*, *PPM1D*, *RAD21*, *SETBP1*, *SF3B1*, *SMC1A*, *SMC3*, *SRSF2*, *TP53*, *U2AF1* and *ZRSR2* (**Supplemental Table 1**).

The presence of CHIP mutations in cardiovascular tissues was studied on DNA level in a subset of 25 confirmed CHIP mutation carriers. Specifically, we looked for respective mutations in atherosclerosis-affected coronary and carotid arteries, and myocardium of the left ventricle. Samples with VAF between 0.5% and 30.0% were analyzed. 18 identical CHIP mutations (i.e. in 72.0% of cases) were identified in at least one corresponding tissue, whereas no *de novo* mutation was detected on tissue level – serving as internal quality control. In 13 (52.0%) atherosclerosis-affected samples of the proximal left anterior descending (LAD) coronary artery CHIP mutations were retrieved: *DNMT3A* (5 mutations), *TET2* (4), *ASXL1* (1), *CBL* (1), *PPM1D* (1), and *SMC3* (1). In atherosclerosis-affected carotid artery samples 10 mutations in *DNMT3A* (6 mutations), *TET2* (3) and *PPM1D* (1) were identified, as well as 4 in the myocardium of the left ventricle of atherosclerosis-affected individuals in *DNMT3A* (1), *TET2* (2) and *SMC3* (1). In general, leukocytes are the only DNA-containing cells in whole blood. In arterial tissue, various cells - e.g. smooth muscle cells, endothelial cells, leukocytes - carry DNA. Given the low proportion of leukocytes in atherosclerotic arterial tissue, an enrichment of CHIP-affected leukocytes - as identified for PPM1D - is likely in plaques (**Table 1**, Supplemental Figure 2).

**Table 1.** This study provides evidence that CHIP-mutated leukocytes have the potential to invade human atherosclerotic plaques in coronary and carotid arteries, and heart muscle from peripheral blood. Out of 25 unique CHIP mutations (identified in whole blood), 18 mutations (shown) were identified in at least one corresponding tissue of interest. Tissue sequencing of CHIP mutation carriers revealed that identical CHIP mutations were identified in the corresponding coronary artery (n=13), carotid artery (n=10) and left ventricular heart muscle (n=4) (right). Provided are mutations on DNA level and variant allele frequency (VAF) in %.

| **CHIP-affected gene**<br>(mutation) | **Whole Blood**<br>(VAF in %) | **Coronary**<br>(VAF in %) | **Carotid**<br>(VAF in %) | **Heart Muscle**<br>(VAF in %) |
| --- | --- | --- | --- | --- |
| **ASXL1** (c.1772dup) | 5.2 | 1.2 | 0 | 0 |
| **CBL** (c.1211G>A) | 9.1 | 2.2 | 0 | 0 |
| **DNMT3A** (c.1628G>C) | 21.6 | 0 | 5.4 | 0 |
| **DNMT3A** (c.976C>T) | 20.5 | 0 | 2.4 | 0 |
| **DNMT3A** (c.2333T>G) | 6.5 | 2.5 | 0 | 0 |
| **DNMT3A** (c.2245C>T) | 5.6 | 1.5 | 0 | 1.5 |
| **DNMT3A** (c.1726_1729delinsC) | 22.6 | 5.9 | 5.8 | 0 |
| **DNMT3A** (c.1969G>A) | 5.3 | 1.0 | 1.1 | 0 |
| **DNMT3A** (c.2204A>G) | 11.3 | 0 | 2.5 | 0 |
| **DNMT3A** (c.2104G>T) | 11.8 | 1.4 | 2.7 | 0 |
| **PPM1D** (c.1535del) | 0.7 | 0 | 2.0 | 0 |
| **PPM1D** (c.1535del) | 0.9 | 1.8 | 0 | 0 |
| **SMC3** (c.3598G>A) | 6.0 | 1.1 | 0 | 1.0 |
| **TET2** (c.2839C>T) | 19.2 | 4.0 | 5.1 | 0 |
| **TET2** (c.4193T>G) | 6.4 | 2.1 | 2.1 | 0 |
| **TET2** (c.1219del) | 27.7 | 4.4 | 0 | 2.0 |
| **TET2** (c.4546C>T) | 29.7 | 5.1 | 0 | 2.6 |
| **TET2** (c.5454_5458del) | 9.8 | 0 | 2.1 | 0 |

### Visualization of specific CHIP mutations

To further detect and visualize specific point mutations of interest on RNA level in single cells custom made mutaFISH^TM^ probes (Abnova, Taiwan) were designed.^25, 26^ The targeted *DNMT3A* mutations c.2245T>C and c.2333G>T, initially identified in whole blood of CAD patients, were identified in leukocytes of the corresponding mutation carriers in human atherosclerotic plaque (**Figure 2**). For optimal orientation, all arterial samples were also stained for hematoxylin and eosin (HE) and Elastin van Gieson (EvG) (**Supplemental Figure 3**). Further, staining for CD68 revealed that CHIP-affected leukocytes in these cases could be identified as macrophages (**Supplemental Figure 4**). CHIP-mutated macrophages were mainly identified in the shoulder regions of advanced and inflammation rich plaques of human coronary and carotid samples (**Figure 2, Supplemental Figure 4<u>+5</u>**).

**Figure 2.**
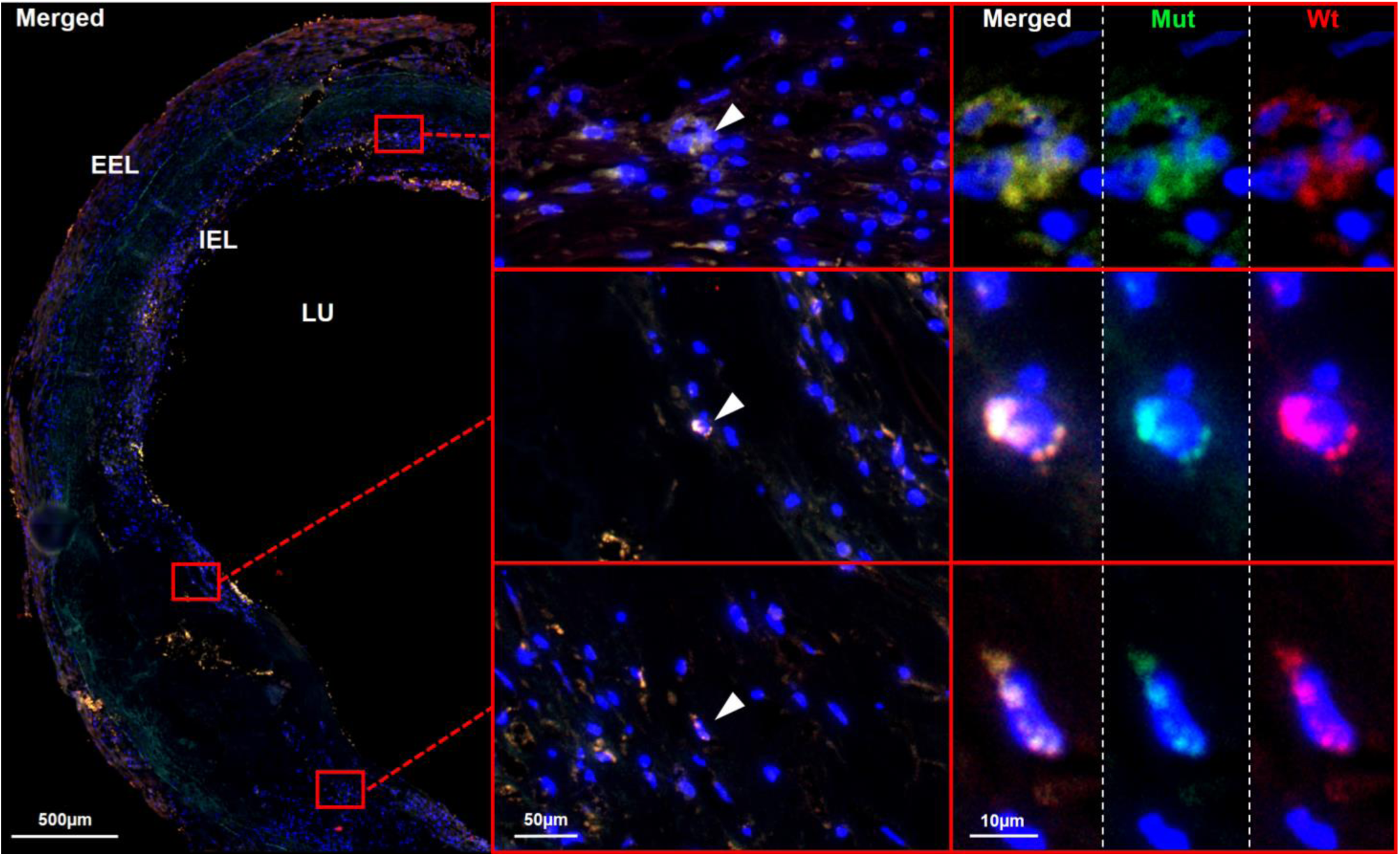
CHIP-mutated cells have the potential to invade from the peripheral blood into human atherosclerotic lesions. mutaFISH™ allowed detection of single nucleotide exchanges at the RNA level in single cells. Staining for a specific DNMT3A mutation (mut) and wild type (wt) at the RNA level was performed *in situ* in advanced atherosclerotic plaques of human FFPE coronary tissue. **Left panel**: merged overview of an advanced atherosclerotic plaque. Red boxes highlight areas of interest enlarged in the following panels. **Middle panel**: CHIP-affected leukocytes were identified in the inflammatory shoulder regions of human atherosclerotic plaque (white arrow). **Right panel**: The DNMTA3 mutation c.2333G>T was detected on single cell resolution via the green signal, the DNMT3A wild type via the red signal and the cell nuclei (DAPI) via the blue signal. DAPI: 4′,6-Diamidin-2-phenylindole; EEL: external elastic lamina; FFPE: formalin-fixed paraffin embedded; IEL: internal elastic lamina; LU: lumen; mutaFISH: mutation-specific Fluorescence In Situ Hybridization.

### CHIP screening by WGS

The 941 patients of the STARNET study undergoing open heart surgery were younger (65.9 years) and WGS sequencing had a lower depth (35-fold) than deep-DNAseq carried out in MISSION.^11^ Overall, 159 specific CHIP mutations in the genes *ASXL1*, *DNMT3A*, *JAK2* or *TET2* were identified in 134 (14.2%) individuals in STARNET (**Supplemental Table 2**). Mean age of these CHIP mutation carriers was 67.1 years and 27 (17.0%) were female. 51 CHIP mutations could be confidentially replicated on RNA level in at least one tissue. Variant analysis in RNAseq data confirmed the CHIP mutations at the RNA level in whole blood (43 mutations), monocyte derived cultured macrophages (10 mutations) and cultured macrophage derived foam cells (5), as well as in atherosclerosis-affected aortic tissue (18) of CAD patients.

### CHIP in macrophages

To study the effects of CHIP mutations on gene expression in macrophages we grouped individuals carrying *TET2* and *ASXL1* mutations and compared these to age- and sex-matched non-mutation carriers (controls). Most macrophages (>80%) belonged to the M0 subtype.

### Differential gene expression analysis of TET2-mutated macrophages

Mean age of *TET2* CHIP mutation carriers (n=3; c.6819G>T; c.6834C>T and c.7698T>C) and matched controls (n=21) showed no significant difference (60.3±12.0 vs 61.8±8.2 years). Detailed information on patient characteristics can be found in **Supplemental Table 3**. In total, 1,523 genes were differentially expressed in *TET2* mutation carriers compared to controls. 1,098 genes were upregulated and 425 genes were downregulated in CHIP mutation carriers. Top upregulated genes were *LINC01882*, *IGKV3-15*, *IGHG2* and *IGHA1*, top downregulated genes were *BMS1P10* and *ZCCHC4* (p_adj._<0.05).

GO analysis revealed as top enriched GO terms T cell receptor complex (GO:0042101), opsonization (GO:0008228) and immune response by circulating immunoglobulin (GO:0002455). In general, immune system and inflammation associated pathways were upregulated (**Figure 3**).

**Figure 3.**
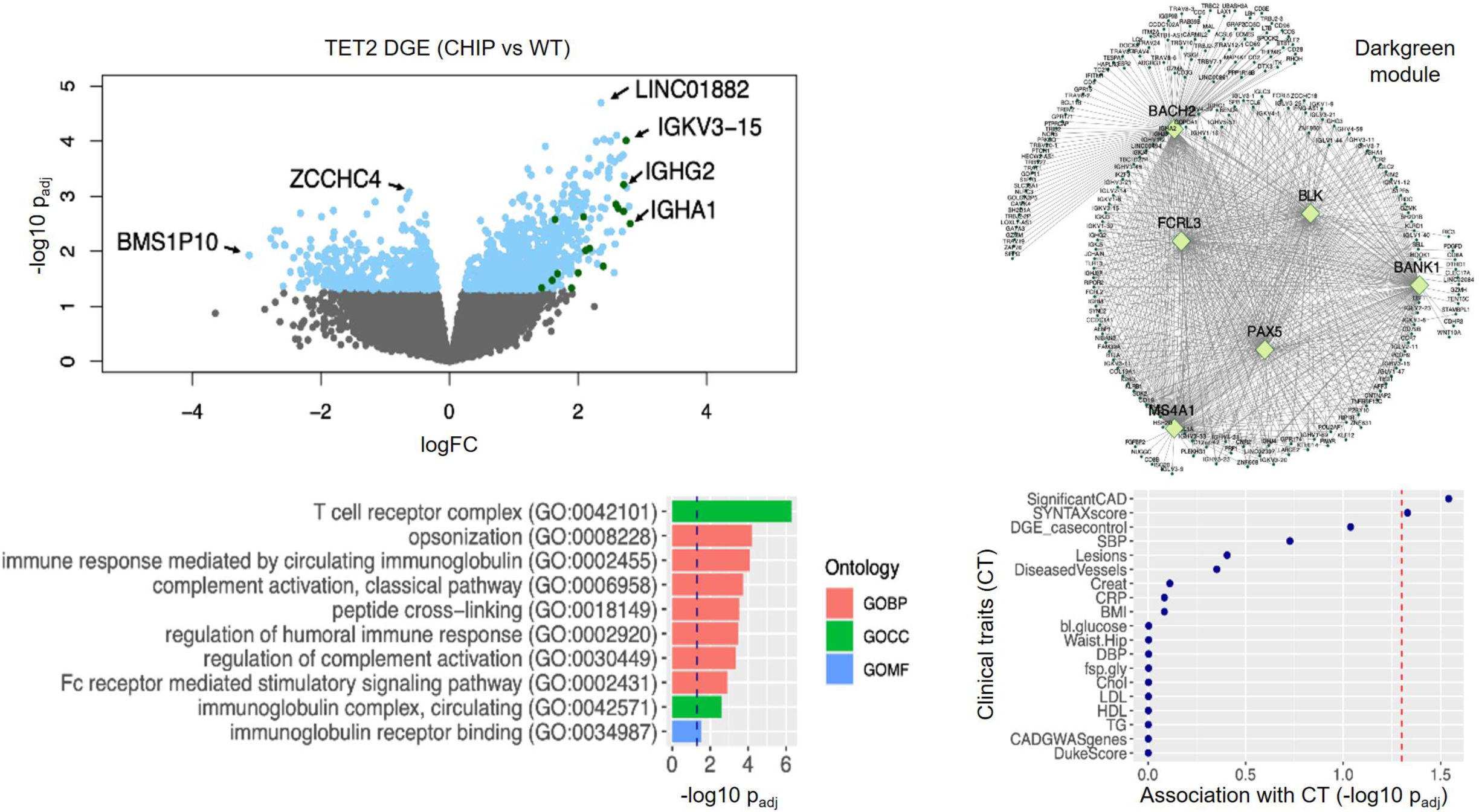
Differentially expressed genes in TET2 CHIP mutation carriers (n=3) versus age and sex matched non-mutation controls (n=21) were enriched in darkgreen module in STARNET macrophage RNAseq data**. Left upper panel**: Volcano plot of differentially expressed genes between TET2 CHIP mutation carriers versus age and sex matched non-mutation controls in STARNET CAD cases. Differentially expressed genes were highlighted in lightblue. Top upregulated and downregulated genes with the smallest p-values or with the biggest log2 fold changes were labeled with gene names. Differentially expressed genes that overlap with darkgreen module genes were highlighted in darkgreen. **Left bottom panel**: Top 10 enriched gene ontologies for differentially expressed genes in TET2 CHIP mutation carriers versus non-mutation controls. Bonferroni corrected p<0.05 was considered statistically significant. **Right upper panel**: Visualization of darkgreen module inferred from STARNET macrophage RNAseq data. Top key drivers were highlighted in yellowgreen. **Right bottom panel**: Dot plot indicating the association of darkgreen module genes with clinical traits. P-value <0.05 (-log10(p) >1.3, cut off was indicated by vertical red line) is considered statistically significant.GO: Gene Ontology; BP: biological pathway; CC: cellular component; MF: molecular function.

Weighted Gene Co-expression Network Analysis (WGCNA) was used to establish co-expression modules based on macrophage RNAseq data from STARNET, to identify functional relations between genes, and based on genetic regulatory networks (GRN) to identify key driver genes. A total of 23 modules were generated from 10,267 transcripts with module sizes ranging between 36 to 2473 genes using the complete macrophage RNAseq data from STARNET. The gene functions of the co-expression modules were analyzed using GO terms. *TET2* mutations in macrophages led to significant perturbation in the darkgreen module (p<0.001), which consists of 38 genes and is primarily involved in immune system related pathways (GO:0019814, GO:0003823, GO:0006959, GO:0006958, GO:0002455, GO:0006956, GO:0002768, GO:0002764, GO:0016064, GO:0019724; all p<0.001). Most relevant key driver genes in this module were *BACH2*, *BANK1*, *BLK*, *FCRL3*, *MS4A1* and *PAX5*. Significant associations were identified for the presence of severity and complexity of CAD based on coronary angiograms, measured by SYNTAX score (**Figure 3**).

### Differential gene expression analysis of ASXL1-mutated macrophages

After matching, mean age of *ASXL1* CHIP mutation carriers (n=3; c.2331C>T; c.4890A>G and c.5137G>A) and controls (n=27) showed no significant difference (67.3±5.5 vs 67.3±5.6 years). Detailed information on patient characteristics can be found in **Supplemental Table 4**. In macrophages of *ASXL1* CHIP mutation carriers 1,222 genes were differentially expressed compared to controls. 665 genes were upregulated and 557 genes were downregulated in CHIP mutation carriers. Top upregulated genes were *HLA-DQB2*, *ASNS* and *SLC25A16*, top downregulated genes were *NPIPB2*, *HNRNPH1* and *POLA2* (p_adj._<0.05). GO analysis revealed intracellular anatomical structure (GO:0005622), RNA binding (GO:0003723) and heterocyclic compound binding (GO:1901363) as top enriched terms. *ASXL1* mutations led to upregulation of cell cycle and metabolic process associated pathways (**Figure 4**).

**Figure 4.**
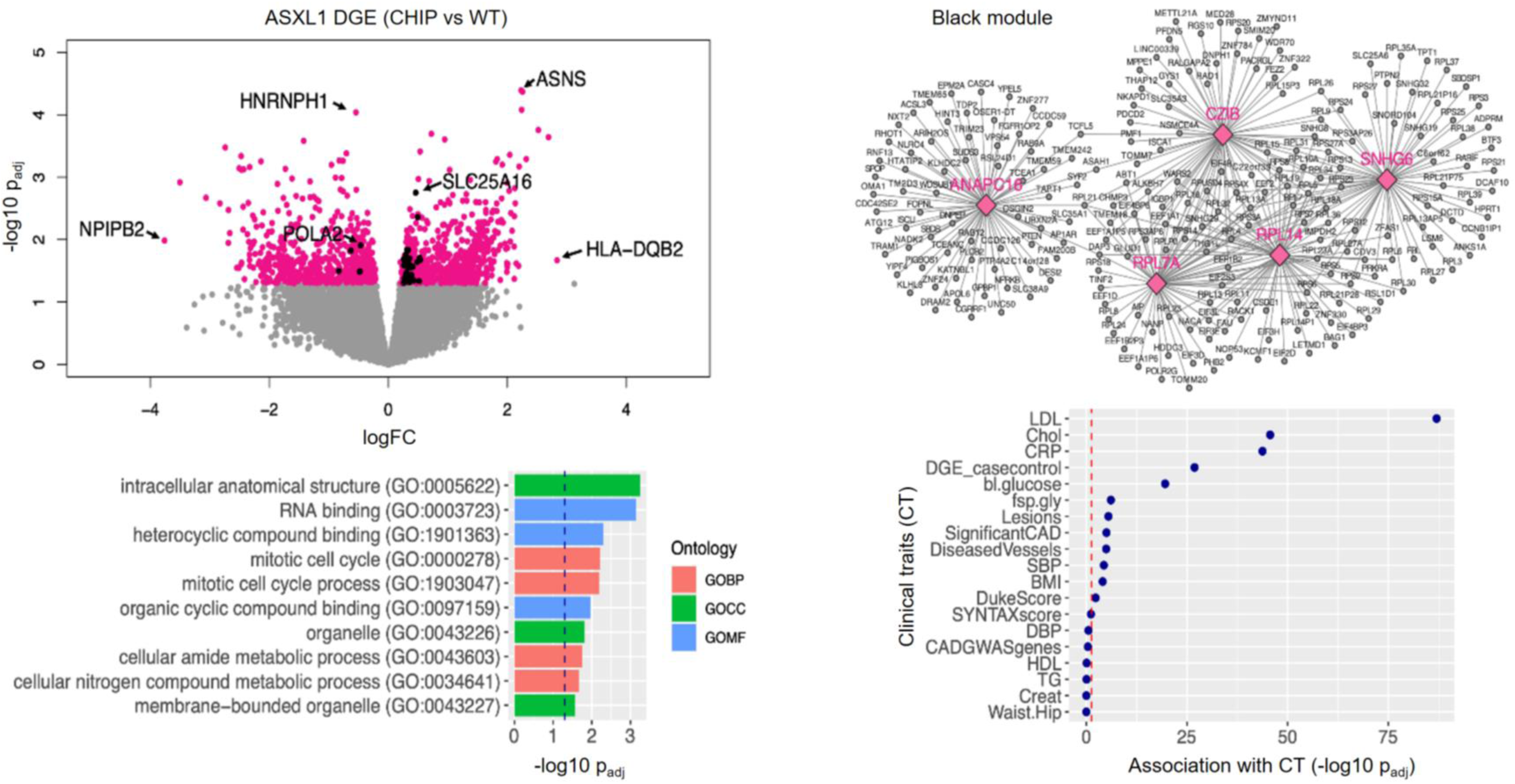
Differentially expressed genes in ASXL1 CHIP mutation carriers (n=3) versus age and sex matched non-mutation controls (n=27) were enriched in black module in STARNET macrophage RNAseq data. **Left upper panel**: Volcano plot of differentially expressed genes (DEGs) between ASXL1 CHIP mutation carriers versus age and sex matched non-mutation controls in STARNET CAD cases. Differentially expressed genes were highlighted in pink. Top upregulated and downregulated genes with the smallest p-values or with the biggest log2 fold changes were labeled with gene names. Differentially expressed genes that overlap with black module genes were highlighted in black. **Left bottom panel**: Top 10 enriched gene ontology (GO) terms for differentially expressed genes in ASXL1 CHIP mutation carriers versus non-mutation controls. Bonferroni corrected p<0.05 was considered statistically significant. **Right upper panel**: Visualization of black module inferred from STARNET macrophage RNAseq data. Top key drivers were highlighted in pink. **Right bottom panel**: Dot plot for the association of black module genes with clinical traits. P-value <0.05 (-log10(p) >1.3, cut off was indicated by vertical red line) is considered statistically significant. GO: Gene Ontology; BP: biological pathway; CC: cellular component; MF: molecular function.

*ASXL1* mutations in macrophages led to significant perturbation in the black module (p<0.001), which consists of 468 genes and is involved in the *intracellular translation of RNA into proteins* (GO:0022626, GO:0006614, GO:0019083, GO:0006613, GO:0045047, GO:0006413, GO:0072599, GO:0000184, GO:0019080, GO:0070972; all p<0.001). Most relevant key driver genes in the black module were *ANAPC16*, *CZIB*, *RPL7A*, *RPL14* and *SNHG6*. The black module was significantly associated with cardiometabolic relevant traits including low-density lipoprotein (LDL) and total cholesterol plasma levels, blood glucose and BMI, inflammation measured by plasma C-reactive protein (CRP) levels, and with the presence and complexity of coronary lesions (**Figure 4**).

## Discussion

To further explore the role of CHIP mutations in CAD, we carried out large-scale, targeted deep-sequencing of genes prone to cause clonal hematopoiesis in MISSION, one of the largest postmortem biobanks focusing on cardiovascular disease. We found at least one characteristic mutation in about 46% of individuals who died with CAD at an average age of 78 years, highlighting that CHIP mutations were significantly more common in CAD patients than previously reported for subjects with unknown CAD status.^2, 5, 27, 28^

In about 70% of affected individuals, the same mutation could be detected within cardiovascular tissues, either in DNA or – in case of coding variants – around one third in RNA. Albeit the cellular composition is different in blood and various tissues, a similar percentage of cells carrying CHIP mutations was found in respective samples. Exemplary visualization of two different *DNMT3A* mutations was achieved within macrophages residing in atherosclerotic human coronary and carotid arteries by in situ fluorescent staining. CHIP mutations, which are thought to promote the development and progression of ischemic heart failure,^6, 29, 30^ were also detected in left ventricular myocardial samples of CAD patients.

The implications of CHIP mutations were then investigated using the STARNET datasets, focusing on CAD patients undergoing open heart surgery (average age 67 years).^11, 31^ Based on whole-genome sequencing and RNAseq data – carried out at a lower depth than the targeted deep-sequencing in MISSION – CHIP mutations were found in about 14% of CAD patients, who most likely represented higher VAFs in affected individuals. Mutations were also detected in aortic samples affected by atherosclerosis, monocyte-derived macrophages, and foam cells. This allowed us to further study the implications of CHIP mutations with respect to clinical presentations and RNA expression patterns.

RNA sequencing revealed large numbers of differentially expressed genes in macrophages of CHIP mutation carriers – as compared to patients free of these mutations. Interestingly, patterns of gene expression also differed between those having mutations in *TET2* and *ASXL1*, i.e. two of the most prominent CHIP mutations.

Patients carrying *TET2-*mutations had a higher CAD burden and – as measured by SYNTAX score – more complex CAD. Macrophages generated *in vitro* by transformation of monocytes from these patients revealed multiple differentially expressed genes. Enrichment analysis revealed upregulation of immune system and inflammation associated pathways. Specifically, *TET2* mutations related to perturbation of a small atherosclerosis relevant macrophage gene expression network with most relevant key driver genes being *BACH2*, *BANK1*, *BLK*, *FCRL3*, *MS4A1* and *PAX5* mainly involved in immune response, cell migration and leukocyte differentiation.

*ASXL1* CHIP mutation carriers revealed upregulation of cardiometabolic relevant traits including low-density lipoprotein (LDL) cholesterol plasma levels, inflammation measured by plasma C-reactive protein (CRP) levels, and – like *TET2* mutations – a higher prevalence and complexity of CAD. In macrophages, *ASXL1* CHIP mutations revealed differentially expressed genes, of note inflammasome related genes were upregulated. Enrichment analysis of differentially expressed genes revealed that *ASXL1* mutations related to perturbations of pathways associated with cell cycle and metabolic processes. Top key drivers affected by CHIP mutations were *ANAPC16*, *CZIB*, *RPL7A*, *RPL14* and *SNHG6* mainly involved in regulation of metabolic processes, ubiquitination and protein synthesis.

Given the relevance of inflammation and cardiovascular conditions that have been associated with CHIP, there appears to be an opportunity to treat selected patients with specific anti-inflammatory strategies,^32–34^ for example, by targeting the *NLRP3* inflammasome, *IL-1β* or *IL-6* with agents like canakinumab (monoclonal antibody against *IL-1β*),^35^ anakinra (an *IL-1β* receptor antagonist),^36–39^ tocilizumab (monoclonal antibody against the *IL-6* receptor),^40, 41^ ziltevekimab (anti-*IL-6* ligand monoclonal antibody),^42^ or colchicine.^43–45^ Interestingly, an exploratory retrospective analysis of the CANTOS trial that studied the effects of canakinumab in patients with an acute coronary syndrome suggested increased inflammatory activation and more cardiovascular events, as well as specific treatment benefits in carriers of CHIP-mutations. However, the value of this analysis is limited as only 40% of the initial CANTOS population was screened for CHIP mutations such that the overall number of CHIP mutation carriers was low.^46^ Our study supports the concept that specific CHIP mutations might identify subgroups of patients with explicit alterations in gene expression in various cardiovascular relevant tissues – mainly stimulating pro-inflammatory mechanisms – which might be relevant in the future for directing specific treatments to these individuals.

### Limitations

Our study has several limitations. Firstly, as we have no longitudinal data, it is unclear how long individual CHIP mutations have been present. CHIP mutations are acquired over decades and their related risks may grow over time. However, the exact time of acquisition of CHIP mutations is difficult to determine and unknown in most cases. Secondly, the limited depth of WGS in STARNET and the focused CHIP screening panel in MISSION may have left mutation carriers undetected. Overlooking CHIP mutation carriers in STARNET may have impaired our sensitivity to discriminate their related effects on clinical and transcriptional characteristics. Thirdly, different DNA sequencing platforms have different levels of coverage, specificity and sensitivity, making it difficult to compare results from different platforms. Standardization of sequencing methods and analysis pipelines would be desirable to ensure comparability of results and improve the accuracy of CHIP screening in the future. Fourth, the relevance of respective mutations and the quantitative burden of VAFs need further clarification. The VAF of CHIP mutations is an essential factor in determining their biological significance, but there is a limited knowledge of optimal VAF ‘cut-offs’ for individual CHIP mutations. Yet, high VAF has been associated with an increased risk of cardiovascular events. In chronic ischemic heart failure (CHF), individual clone size thresholds of less than 2% in *DNMT3A* and *TET2* have already been associated with worse outcomes.^29^ It remains to be clarified which VAF threshold is relevant for individual CHIP mutations and specific phenotypes, and whether these could serve as novel biomarkers or support treatment decisions. Fifth, the relevance of respective mutations in one or the other gene, including the additive effects of multiple mutations needs further investigation. An essential key for future personalized therapy approaches will be to validate these CHIP related outcomes in well-powered, prospective RCTs. Finally, the local effects of individual CHIP-mutated macrophages residing within cardiovascular tissues needs spatial investigation.

## Conclusions

Our data highlight that blood derived circulating CHIP-affected leukocytes have the potential to invade human atherosclerotic lesions in coronary and carotid arteries, aorta, and heart muscle. We visualized CHIP-mutated leukocytes with single cell resolution in human atherosclerotic plaques and staining confirmed these leukocytes as CD68^+^ macrophages. Since these acquired CHIP mutations originate from hematopoietic stem or progenitor cells in the bone marrow and were identified in whole blood, the visualized CHIP mutations cannot originate from resident arterial macrophages. Further, RNA sequencing identified CHIP specific signatures in macrophages, novel key regulators and associations with clinical traits like burden and complexity of CAD (**Figure 1**). Future studies are needed to further characterize underlying molecular mechanisms by which individual mutations contribute to CVDs and whether targeting specific pathways may be valuable for precision medicine in patients carrying mutations encoding for CHIP.

## Novelty and Significance

**What is known? (2-3 bullet points)**

- CHIP represents an independent risk factor for the onset and progression of cardiovascular diseases and death
- CHIP mutations are associated with inflammatory activation of circulating monocytes and T-cells
- Individual CHIP mutations could be important as biomarkers in the future and help identify high-risk patients suitable for anti-inflammatory therapy

**What new information does this article contribute? (max. 200 words)**

Our work provides insights on three open questions in the field. Firstly, we provide higher estimates of the proportion of CAD patients affected by CHIP mutations, if these are studied by focused deep-sequencing in whole blood. Next, we show that CHIP-mutated leukocytes have the potential to invade human plaques and thereby contribute locally to atherosclerosis. Specifically, we visualized macrophages at single cell resolution by a specific *DNMT3A* CHIP mutations probe in coronary plaques. We also identified blood derived CHIP mutations in human coronary and carotid arteries and heart muscle on DNA level. Additionally, we describe previously unknown key-regulatory genes and pro-atherosclerotic alterations at the RNA level of CHIP mutated macrophages. Importantly, different CHIP mutations appear to affect different pathways, networks and modules, which nevertheless all have relevance for CAD progression. These novel results (**Figure 1**) appear to be of relevance for future personalized medicine approaches specifically designed for CHIP mutation carriers.

## Supporting information

CHIP_Supplement

## Acknowledgments

M.v.S drafted the manuscript, designed and coordinated the study. J.K., M.G. and C.B. provided human cardiovascular tissues in MISSION. M.v.S, S.B., J.F., D.B. and L.O. performed the CHIP screening in MISSION. S.B. performed the mutaFISH^TM^ experiment. A.M., K.H., A.R., J.C.K. and J.L.M.B. contributed data from STARNET. Y.W., C.J.H., S.K.B.G., M.M., H.G., K.K., A.S., C.M., N.L., J.L.M.B. and H.S. provided relevant intellectual input within the Leducq PlaqOmics Consortium. B.V., T.K., Z.C., A.M., H.B.S. and L.M. were involved RNAseq analyses. J.S.H., F.B. and W.K. provided relevant intellectual input regarding hematological aspects and biomarkers. A.M.Z. and S.D. supported within the joint DZHK CHIP moonshot project. All authors critically reviewed the manuscript and agreed with the final version of the manuscript, written by M.v.S. J.L.M.B. and H.S.. Special thanks for scientific support to the Munich Leukemia Laboratory (MLL), Germany. Illustrations contain image material available at Servier Medical Art under a creative commons attribution 3.0 unported license.

## Supplement

**Supplemental Table 1** – List of CHIP mutations in MISSION

**Supplemental Table 2** – List of CHIP mutations in STARNET

**Supplemental Table 3** – STARNET patient characteristics TET2 macrophages

**Supplemental Table 4** – STARNET patient characteristics ASXL1 macrophages

**Supplemental Figure 1** – VAF and distribution of CHIP mutations in MISSION

**Supplemental Figure 2** – Deep-DNAseq identifies CHIP mutations in atherosclerotic coronary and carotid samples, and left ventricular myocardium

**Supplemental Figure 3** – Overview plaque of interest – different stainings

**Supplemental Figure 4** – Visualization of CD68^+^ CHIP mutated macrophage

**Supplemental Figure 5** – DNMT3A CHIP mutation (c.2245C>T) in human atherosclerotic plaque

**Protocol** – adapted mutaFISH^TM^ protocol

## References

1. Genovese G, Kahler AK, Handsaker RE, Lindberg J, Rose SA, Bakhoum SF, Chambert K, Mick E, Neale BM, Fromer M, Purcell SM, Svantesson O, Landen M, Hoglund M, Lehmann S, Gabriel SB, Moran JL, Lander ES, Sullivan PF, Sklar P, Gronberg H, Hultman CM and McCarroll SA. Clonal hematopoiesis and blood-cancer risk inferred from blood DNA sequence. N Engl J Med. 2014;371:2477–87.

2. Jaiswal S, Fontanillas P, Flannick J, Manning A, Grauman PV, Mar BG, Lindsley RC, Mermel CH, Burtt N, Chavez A, Higgins JM, Moltchanov V, Kuo FC, Kluk MJ, Henderson B, Kinnunen L, Koistinen HA, Ladenvall C, Getz G, Correa A, Banahan BF, Gabriel S, Kathiresan S, Stringham HM, McCarthy MI, Boehnke M, Tuomilehto J, Haiman C, Groop L, Atzmon G, Wilson JG, Neuberg D, Altshuler D and Ebert BL. Age-related clonal hematopoiesis associated with adverse outcomes. N Engl J Med. 2014;371:2488–98.

3. Young AL, Challen GA, Birmann BM and Druley TE. Clonal haematopoiesis harbouring AML-associated mutations is ubiquitous in healthy adults. Nat Commun. 2016;7:12484.

4. Hecker JS, Hartmann L, Riviere J, Buck MC, van der Garde M, Rothenberg-Thurley M, Fischer L, Winter S, Ksienzyk B, Ziemann F, Solovey M, Rauner M, Tsourdi E, Sockel K, Schneider M, Kubasch AS, Nolde M, Hausmann D, Paulus AC, Lutzner J, Roth A, Bassermann F, Spiekermann K, Marr C, Hofbauer LC, Platzbecker U, Metzeler KH and Gotze KS. CHIP and hips: clonal hematopoiesis is common in patients undergoing hip arthroplasty and is associated with autoimmune disease. Blood. 2021;138:1727–1732.

5. Jaiswal S, Natarajan P, Silver AJ, Gibson CJ, Bick AG, Shvartz E, McConkey M, Gupta N, Gabriel S, Ardissino D, Baber U, Mehran R, Fuster V, Danesh J, Frossard P, Saleheen D, Melander O, Sukhova GK, Neuberg D, Libby P, Kathiresan S and Ebert BL. Clonal Hematopoiesis and Risk of Atherosclerotic Cardiovascular Disease. N Engl J Med. 2017;377:111–121.

6. Dorsheimer L, Assmus B, Rasper T, Ortmann CA, Ecke A, Abou-El-Ardat K, Schmid T, Brune B, Wagner S, Serve H, Hoffmann J, Seeger F, Dimmeler S, Zeiher AM and Rieger MA. Association of Mutations Contributing to Clonal Hematopoiesis With Prognosis in Chronic Ischemic Heart Failure. JAMA Cardiol. 2019;4:25–33.

7. Bhattacharya R, Zekavat SM, Haessler J, Fornage M, Raffield L, Uddin MM, Bick AG, Niroula A, Yu B, Gibson C, Griffin G, Morrison AC, Psaty BM, Longstreth WT, Bis JC, Rich SS, Rotter JI, Tracy RP, Correa A, Seshadri S, Johnson A, Collins JM, Hayden KM, Madsen TE, Ballantyne CM, Jaiswal S, Ebert BL, Kooperberg C, Manson JE, Whitsel EA, Program NT-OfPM, Natarajan P and Reiner AP. Clonal Hematopoiesis Is Associated With Higher Risk of Stroke. Stroke. 2022;53:788–797.

8. Gumuser ED, Schuermans A, Cho SMJ, Sporn ZA, Uddin MM, Paruchuri K, Nakao T, Yu Z, Haidermota S, Hornsby W, Weeks LD, Niroula A, Jaiswal S, Libby P, Ebert BL, Bick AG, Natarajan P and Honigberg MC. Clonal Hematopoiesis of Indeterminate Potential Predicts Adverse Outcomes in Patients With Atherosclerotic Cardiovascular Disease. J Am Coll Cardiol. 2023;81:1996–2009.

9. Fuster JJ, MacLauchlan S, Zuriaga MA, Polackal MN, Ostriker AC, Chakraborty R, Wu CL, Sano S, Muralidharan S, Rius C, Vuong J, Jacob S, Muralidhar V, Robertson AA, Cooper MA, Andres V, Hirschi KK, Martin KA and Walsh K. Clonal hematopoiesis associated with TET2 deficiency accelerates atherosclerosis development in mice. Science. 2017;355:842–847.

10. Libby P and Ebert BL. CHIP (Clonal Hematopoiesis of Indeterminate Potential): Potent and Newly Recognized Contributor to Cardiovascular Risk. Circulation. 2018;138:666–668.

11. Koplev S, Seldin S, Sukhavasi K, Ermel R, Pang S, Zeng L, Bankier S, Di Narzo A, Cheng H, Meda V, Ma A, Talukdar H, Cohain A, Amadori L, Argmann C, Houten S, Franzén O, Mocci G, Meelu O, Ishikawa K, Whatling C, Jain A, Jain R, Gan L, Giannarelli C, Roussos P, Hao K, Schunkert H, Michoel T, Ruusalepp A, Schadt E, Kovacic J, Lusis A and Björkegren J. A mechanistic framework for cardiometabolic and coronary artery diseases. Nature Cardiovascular Research. 2022;1:85–100.

12. Li H and Durbin R. Fast and accurate short read alignment with Burrows-Wheeler transform. Bioinformatics. 2009;25:1754–60.

13. Li H and Durbin R. Fast and accurate long-read alignment with Burrows-Wheeler transform. Bioinformatics. 2010;26:589–95.

14. Nowak J, Koscinska K, Mika-Witkowska R, Rogatko-Koros M, Mizia S, Jaskula E, Polak M, Mordak-Domagala M, Lange J, Gronkowska A, Jedrzejczak WW, Kyrcz-Krzemien S, Markiewicz M, Dzierzak-Mietla M, Tomaszewska A, Nasilowska-Adamska B, Szczepinski A, Halaburda K, Hellmann A, Komarnicki M, Gil L, Czyz A, Wachowiak J, Baranska M, Kowalczyk J, Drabko K, Gozdzik J, Wysoczanska B, Bogunia-Kubik K, Graczyk-Pol E, Witkowska A, Marosz-Rudnicka A, Nestorowicz K, Dziopa J, Szlendak U, Warzocha K and Lange AA. Donor NK cell licensing in control of malignancy in hematopoietic stem cell transplant recipients. Am J Hematol. 2014;89:E176–83.

15. Broadinstitute. Picard. 2022;2022:Picard v1.118.

16. McKenna A, Hanna M, Banks E, Sivachenko A, Cibulskis K, Kernytsky A, Garimella K, Altshuler D, Gabriel S, Daly M and DePristo MA. The Genome Analysis Toolkit: a MapReduce framework for analyzing next-generation DNA sequencing data. Genome Res. 2010;20:1297–303.

17. Poplin R, Ruano-Rubio V, DePristo MA, Fennell TJ, Carneiro MO, Van der Auwera GA, Kling DE, Gauthier LD, Levy-Moonshine A, Roazen D, Shakir K, Thibault J, Chandran S, Whelan C, Lek M, Gabriel S, Daly MJ, Neale B, MacArthur DG and Banks E. Scaling accurate genetic variant discovery to tens of thousands of samples. bioRxiv. 2018:201178.

18. DePristo MA, Banks E, Poplin R, Garimella KV, Maguire JR, Hartl C, Philippakis AA, del Angel G, Rivas MA, Hanna M, McKenna A, Fennell TJ, Kernytsky AM, Sivachenko AY, Cibulskis K, Gabriel SB, Altshuler D and Daly MJ. A framework for variation discovery and genotyping using next-generation DNA sequencing data. Nat Genet. 2011;43:491–8.

19. Van der Auwera GA, Carneiro MO, Hartl C, Poplin R, Del Angel G, Levy-Moonshine A, Jordan T, Shakir K, Roazen D, Thibault J, Banks E, Garimella KV, Altshuler D, Gabriel S and DePristo MA. From FastQ data to high confidence variant calls: the Genome Analysis Toolkit best practices pipeline. Curr Protoc Bioinformatics. 2013;43:11 10 1–11 10 33.

20. Huynh-Thu VA and Geurts P. dynGENIE3: dynamical GENIE3 for the inference of gene networks from time series expression data. Sci Rep. 2018;8:3384.

21. Lizio M, Harshbarger J, Shimoji H, Severin J, Kasukawa T, Sahin S, Abugessaisa I, Fukuda S, Hori F, Ishikawa-Kato S, Mungall CJ, Arner E, Baillie JK, Bertin N, Bono H, de Hoon M, Diehl AD, Dimont E, Freeman TC, Fujieda K, Hide W, Kaliyaperumal R, Katayama T, Lassmann T, Meehan TF, Nishikata K, Ono H, Rehli M, Sandelin A, Schultes EA, t Hoen PA, Tatum Z, Thompson M, Toyoda T, Wright DW, Daub CO, Itoh M, Carninci P, Hayashizaki Y, Forrest AR, Kawaji H and consortium F. Gateways to the FANTOM5 promoter level mammalian expression atlas. Genome Biol. 2015;16:22.

22. Abugessaisa I, Ramilowski JA, Lizio M, Severin J, Hasegawa A, Harshbarger J, Kondo A, Noguchi S, Yip CW, Ooi JLC, Tagami M, Hori F, Agrawal S, Hon CC, Cardon M, Ikeda S, Ono H, Bono H, Kato M, Hashimoto K, Bonetti A, Kato M, Kobayashi N, Shin J, de Hoon M, Hayashizaki Y, Carninci P, Kawaji H and Kasukawa T. FANTOM enters 20th year: expansion of transcriptomic atlases and functional annotation of non-coding RNAs. Nucleic Acids Res. 2021;49:D892–D898.

23. Shu L, Zhao Y, Kurt Z, Byars SG, Tukiainen T, Kettunen J, Orozco LD, Pellegrini M, Lusis AJ, Ripatti S, Zhang B, Inouye M, Makinen VP and Yang X. Mergeomics: multidimensional data integration to identify pathogenic perturbations to biological systems. BMC Genomics. 2016;17:874.

24. Ding J, Blencowe M, Nghiem T, Ha SM, Chen YW, Li G and Yang X. Mergeomics 2.0: a web server for multi-omics data integration to elucidate disease networks and predict therapeutics. Nucleic Acids Res. 2021;49:W375–W387.

25. Larsson C, Grundberg I, Soderberg O and Nilsson M. In situ detection and genotyping of individual mRNA molecules. Nat Methods. 2010;7:395–7.

26. Larsson C, Koch J, Nygren A, Janssen G, Raap AK, Landegren U and Nilsson M. In situ genotyping individual DNA molecules by target-primed rolling-circle amplification of padlock probes. Nat Methods. 2004;1:227–32.

27. Yu Z, Fidler TP, Ruan Y, Vlasschaert C, Nakao T, Uddin MM, Mack T, Niroula A, Heimlich JB, Zekavat SM, Gibson CJ, Griffin GK, Wang Y, Peloso GM, Heard-Costa N, Levy D, Vasan RS, Aguet F, Ardlie K, Taylor KD, Rich SS, Rotter JI, Libby P, Jaiswal S, Ebert BL, Bick AG, Tall AR and Natarajan P. Genetic modification of inflammation and clonal hematopoiesis-associated coronary artery disease. medRxiv. 2022:2022.12.08.22283237.

28. Jaiswal S and Libby P. Clonal haematopoiesis: connecting ageing and inflammation in cardiovascular disease. Nat Rev Cardiol. 2020;17:137–144.

29. Assmus B, Cremer S, Kirschbaum K, Culmann D, Kiefer K, Dorsheimer L, Rasper T, Abou-El-Ardat K, Herrmann E, Berkowitsch A, Hoffmann J, Seeger F, Mas-Peiro S, Rieger MA, Dimmeler S and Zeiher AM. Clonal haematopoiesis in chronic ischaemic heart failure: prognostic role of clone size for DNMT3A- and TET2-driver gene mutations. Eur Heart J. 2021;42:257–265.

30. Pascual-Figal DA, Bayes-Genis A, Diez-Diez M, Hernandez-Vicente A, Vazquez-Andres D, de la Barrera J, Vazquez E, Quintas A, Zuriaga MA, Asensio-Lopez MC, Dopazo A, Sanchez-Cabo F and Fuster JJ. Clonal Hematopoiesis and Risk of Progression of Heart Failure With Reduced Left Ventricular Ejection Fraction. J Am Coll Cardiol. 2021;77:1747–1759.

31. Franzen O, Ermel R, Cohain A, Akers NK, Di Narzo A, Talukdar HA, Foroughi-Asl H, Giambartolomei C, Fullard JF, Sukhavasi K, Koks S, Gan LM, Giannarelli C, Kovacic JC, Betsholtz C, Losic B, Michoel T, Hao K, Roussos P, Skogsberg J, Ruusalepp A, Schadt EE and Bjorkegren JL. Cardiometabolic risk loci share downstream cis- and trans-gene regulation across tissues and diseases. Science. 2016;353:827–30.

32. Tall AR and Bornfeldt KE. Inflammasomes and Atherosclerosis: a Mixed Picture. Circ Res. 2023;132:1505–1520.

33. Hettwer J, Hinterdobler J, Miritsch B, Deutsch MA, Li X, Mauersberger C, Moggio A, Braster Q, Gram H, Robertson AAB, Cooper MA, Gross O, Krane M, Weber C, Koenig W, Soehnlein O, Adamstein NH, Ridker P, Schunkert H, Libby P, Kessler T and Sager HB. Interleukin-1beta suppression dampens inflammatory leucocyte production and uptake in atherosclerosis. Cardiovasc Res. 2022;118:2778–2791.

34. Abbate A, Toldo S, Marchetti C, Kron J, Van Tassell BW and Dinarello CA. Interleukin-1 and the Inflammasome as Therapeutic Targets in Cardiovascular Disease. Circ Res. 2020;126:1260–1280.

35. Ridker PM, Everett BM, Thuren T, MacFadyen JG, Chang WH, Ballantyne C, Fonseca F, Nicolau J, Koenig W, Anker SD, Kastelein JJP, Cornel JH, Pais P, Pella D, Genest J, Cifkova R, Lorenzatti A, Forster T, Kobalava Z, Vida-Simiti L, Flather M, Shimokawa H, Ogawa H, Dellborg M, Rossi PRF, Troquay RPT, Libby P, Glynn RJ and Group CT. Antiinflammatory Therapy with Canakinumab for Atherosclerotic Disease. N Engl J Med. 2017;377:1119–1131.

36. Abbate A, Trankle CR, Buckley LF, Lipinski MJ, Appleton D, Kadariya D, Canada JM, Carbone S, Roberts CS, Abouzaki N, Melchior R, Christopher S, Turlington J, Mueller G, Garnett J, Thomas C, Markley R, Wohlford GF, Puckett L, Medina de Chazal H, Chiabrando JG, Bressi E, Del Buono MG, Schatz A, Vo C, Dixon DL, Biondi-Zoccai GG, Kontos MC and Van Tassell BW. Interleukin-1 Blockade Inhibits the Acute Inflammatory Response in Patients With ST-Segment-Elevation Myocardial Infarction. J Am Heart Assoc. 2020;9:e014941.

37. Buckley LF and Abbate A. Interleukin-1 blockade in cardiovascular diseases: a clinical update. Eur Heart J. 2018;39:2063–2069.

38. Dragoljevic D, Lee MKS, Louis C, Shihata W, Kraakman MJ, Hansen J, Masters SL, Hanaoka BY, Nagareddy PR, Lancaster GI, Wicks IP and Murphy AJ. Inhibition of interleukin-1beta signalling promotes atherosclerotic lesion remodelling in mice with inflammatory arthritis. Clin Transl Immunology. 2020;9:e1206.

39. Ku EJ, Kim BR, Lee JI, Lee YK, Oh TJ, Jang HC and Choi SH. The Anti-Atherosclerosis Effect of Anakinra, a Recombinant Human Interleukin-1 Receptor Antagonist, in Apolipoprotein E Knockout Mice. Int J Mol Sci. 2022;23.

40. Hartman J and Frishman WH. Inflammation and atherosclerosis: a review of the role of interleukin-6 in the development of atherosclerosis and the potential for targeted drug therapy. Cardiol Rev. 2014;22:147–51.

41. Poznyak AV, Bharadwaj D, Prasad G, Grechko AV, Sazonova MA and Orekhov AN. Anti-Inflammatory Therapy for Atherosclerosis: Focusing on Cytokines. Int J Mol Sci. 2021;22.

42. Ridker PM, Devalaraja M, Baeres FMM, Engelmann MDM, Hovingh GK, Ivkovic M, Lo L, Kling D, Pergola P, Raj D, Libby P, Davidson M and Investigators R. IL-6 inhibition with ziltivekimab in patients at high atherosclerotic risk (RESCUE): a double-blind, randomised, placebo-controlled, phase 2 trial. Lancet. 2021;397:2060–2069.

43. Tardif JC, Kouz S, Waters DD, Bertrand OF, Diaz R, Maggioni AP, Pinto FJ, Ibrahim R, Gamra H, Kiwan GS, Berry C, Lopez-Sendon J, Ostadal P, Koenig W, Angoulvant D, Gregoire JC, Lavoie MA, Dube MP, Rhainds D, Provencher M, Blondeau L, Orfanos A, L’Allier PL, Guertin MC and Roubille F. Efficacy and Safety of Low-Dose Colchicine after Myocardial Infarction. N Engl J Med. 2019;381:2497–2505.

44. Nidorf SM, Fiolet ATL, Mosterd A, Eikelboom JW, Schut A, Opstal TSJ, The SHK, Xu XF, Ireland MA, Lenderink T, Latchem D, Hoogslag P, Jerzewski A, Nierop P, Whelan A, Hendriks R, Swart H, Schaap J, Kuijper AFM, van Hessen MWJ, Saklani P, Tan I, Thompson AG, Morton A, Judkins C, Bax WA, Dirksen M, Alings M, Hankey GJ, Budgeon CA, Tijssen JGP, Cornel JH, Thompson PL and LoDoCo2 Trial I. Colchicine in Patients with Chronic Coronary Disease. N Engl J Med. 2020;383:1838–1847.

45. Opstal TSJ, van Broekhoven A, Fiolet ATL, Mosterd A, Eikelboom JW, Nidorf SM, Thompson PL, Budgeon CA, Bartels L, de Nooijer R, Bax WA, Tijssen JGP, El Messaoudi S and Cornel JH. Long-Term Efficacy of Colchicine in Patients With Chronic Coronary Disease: Insights From LoDoCo2. Circulation. 2022;145:626–628.

46. Svensson EC, Madar A, Campbell CD, He Y, Sultan M, Healey ML, Xu H, D’Aco K, Fernandez A, Wache-Mainier C, Libby P, Ridker PM, Beste MT and Basson CT. TET2-Driven Clonal Hematopoiesis and Response to Canakinumab: An Exploratory Analysis of the CANTOS Randomized Clinical Trial. JAMA Cardiol. 2022;7:521–528.

