## Supplementary material for "Leukocytes carrying *Clonal Hematopoiesis of Indeterminate Potential* (CHIP) Mutations invade Human Atherosclerotic Plaques": CHIP_Supplement

von Scheidt et al.

**Content**

**Supplemental Table 1** – List of CHIP mutations in MISSION

**Supplemental Table 2** – List of CHIP mutations in STARNET

**Supplemental Table 3** – STARNET patient characteristics TET2 macrophages

**Supplemental Table 4** – STARNET patient characteristics ASXL1 macrophages

**Supplemental Figure 1** – VAF and distribution of CHIP mutations in MISSION

**Supplemental Figure 2** – DeepDNAseq identifies CHIP mutations in atherosclerotic coronary and carotid samples, and left ventricular myocardium

**Supplemental Figure 3** – Overview plaque of interest – different stainings

**Supplemental Figure 4** – Visualization of $CD68^+$ CHIP mutated macrophage

**Supplemental Figure 5** – DNMT3A CHIP mutation (c.2245C>T) in human atherosclerotic plaque

**Protocol** – Adapted mutaFISH$^{TM}$ protocol

**Supplemental Table 1** – List of 445 unique CHIP mutations identified in MISSION based on deep-DNA-sequencing. Provided are gene name, confirmation of CHIP mutation – polymorphisms, variants, synonymous and uncertain mutations were excluded, change on DNA level, change on amino acid (AA) level and variant allele frequency (VAF).

| Gene | CHIP result | DNA result | AA result | VAF (%) |
|---|---|---|---|---|
| ASXL1 | mutated | c.1534C>T | p.Gln512* | 4.1 |
| ASXL1 | mutated | c.1564C>T | p.Gln522* | 4.7 |
| ASXL1 | mutated | c.1585C>T | p.Gln529* | 16.3 |
| ASXL1 | mutated | c.1720-2A>G | p.splice site mutation | 6.3 |
| ASXL1 | mutated | c.1749G>A | p.Trp583* | 31.5 |
| ASXL1 | mutated | c.1762C>T | p.Gln588* | 5.5 |
| ASXL1 | mutated | c.1772dup | p.Tyr591* | 5.2 |
| ASXL1 | mutated | c.1772dup | p.Tyr591* | 3.0 |
| ASXL1 | mutated | c.1900_1922del | p.Glu635Argfs*15 | 7.6 |
| ASXL1 | mutated | c.1900_1922del | p.Glu635Argfs*15 | 6.6 |
| ASXL1 | mutated | c.1900_1922del | p.Glu635Argfs*15 | 1.5 |
| ASXL1 | mutated | c.1900_1922del | p.Glu635Argfs*15 | 11.0 |
| ASXL1 | mutated | c.1900_1922del | p.Glu635Argfs*15 | 19.6 |
| ASXL1 | mutated | c.1934dup | p.Gly646Trpfs*12 | 18.6 |
| ASXL1 | mutated | c.1934dup | p.Gly646Trpfs*12 | 16.6 |
| ASXL1 | mutated | c.1934dup | p.Gly646Trpfs*12 | 12.4 |
| ASXL1 | mutated | c.1934dup | p.Gly646Trpfs*12 | 10.8 |
| ASXL1 | mutated | c.1934dup | p.Gly646Trpfs*12 | 7.0 |
| ASXL1 | mutated | c.1934dup | p.Gly646Trpfs*12 | 11.2 |
| ASXL1 | mutated | c.1934dup | p.Gly646Trpfs*12 | 16.0 |
| ASXL1 | mutated | c.2069_2075del | p.Asp690Glyfs*11 | 1.5 |
| ASXL1 | mutated | c.2077C>T | p.Arg693* | 2.1 |
| ASXL1 | mutated | c.2083C>T | p.Gln695* | 2.6 |
| ASXL1 | mutated | c.2290del | p.Leu764Tyrfs*8 | 5.5 |
| ASXL1 | mutated | c.2302C>T | p.Gln768* | 22.6 |
| ASXL1 | mutated | c.2324T>G | p.Leu775* | 14.1 |
| ASXL1 | mutated | c.2387G>A | p.Trp796* | 16.3 |
| ASXL1 | mutated | c.2468del | p.Leu823* | 6.1 |
| ASXL1 | mutated | c.2528_2529insCT | p.Thr844* | 2.2 |
| ASXL1 | mutated | c.2676del | p.Asn893Thrfs*15 | 9.6 |
| ASXL1 | mutated | c.2989G>T | p.Glu997* | 7.8 |
| ASXL1 | mutated | c.3554dup | p.Thr1186Hisfs*7 | 2.7 |
| BCOR | mutated | c.4616dup | p.Asn1540Glufs*16 | 7.0 |
| CALR | mutated | c.1154_1155insTTGTC | p.Lys385Asnfs*47 | 28.8 |
| CBL | mutated | c.1009del | p.Tyr337Ilefs*13 | 5.0 |
| CBL | mutated | c.1102T>C | p.Tyr368His | 11.5 |
| CBL | mutated | c.1129A>G | p.Thr377Ala | 8.0 |
| CBL | mutated | c.1139T>C | p.Leu380Pro | 1.7 |
| CBL | mutated | c.1145A>G | p.Lys382Arg | 4.3 |
| CBL | mutated | c.1175A>G | p.Lys392Arg | 1.3 |

| | | | | |
|---|---|---|---|---|
| CBL | mutated | c.1211G>A | p.Cys404Tyr | 29.2 |
| CBL | mutated | c.1211G>A | p.Cys404Tyr | 9.1 |
| CBL | mutated | c.1211G>A | p.Cys404Tyr | 3.0 |
| CBL | mutated | c.1211G>A | p.Cys404Tyr | 4.2 |
| CBL | mutated | c.1254C>A | p.Phe418Leu | 6.7 |
| CBL | mutated | c.1268T>A | p.Ile423Asn | 23.0 |
| CBL | mutated | c.1694del | p.Leu565Cysfs*50 | 3.5 |
| DNMT3A | mutated | c.1014+1G>T | p.splice site mutation | 4.0 |
| DNMT3A | mutated | c.1021_1022del | p.Val341* | 1.9 |
| DNMT3A | mutated | c.1040T>C | p.Leu347Pro | 9.6 |
| DNMT3A | mutated | c.1058_1066del | p.Ala353_Gln356delinsGlu | 11.0 |
| DNMT3A | mutated | c.1069_1086dup | p.Ala357_Gln362dup | 1.2 |
| DNMT3A | mutated | c.1077_1078dup | p.Asn360Thrfs*48 | 2.1 |
| DNMT3A | mutated | c.1136G>A | p.Arg379His | 3.3 |
| DNMT3A | mutated | c.1138_1147del | p.Ala380Cysfs*24 | 1.8 |
| DNMT3A | mutated | c.1152_1155del | p.Phe384Leufs*22 | 8.0 |
| DNMT3A | mutated | c.1156del | p.Val386Cysfs*21 | 3.0 |
| DNMT3A | mutated | c.1223_1226del | p.Glu408Glyfs*242 | 5.1 |
| DNMT3A | mutated | c.1226G>A | p.Trp409* | 1.7 |
| DNMT3A | mutated | c.1229C>T | p.Ala410Val | 5.5 |
| DNMT3A | mutated | c.1234_1235insA | p.Gly412Glufs*9 | 1.1 |
| DNMT3A | mutated | c.1238dup | p.Phe414Leufs*7 | 2.7 |
| DNMT3A | mutated | c.1410del | p.Ile471Leufs*180 | 1.4 |
| DNMT3A | mutated | c.1428del | p.Glu477Serfs*174 | 1.5 |
| DNMT3A | mutated | c.1429G>C | p.Glu477Gln | 5.5 |
| DNMT3A | mutated | c.1481G>A | p.Cys494Tyr | 2.9 |
| DNMT3A | mutated | c.1489T>C | p.Cys497Arg | 2.2 |
| DNMT3A | mutated | c.1498del | p.Leu500Serfs*151 | 2.3 |
| DNMT3A | mutated | c.1507dup | p.Thr503Asnfs*43 | 14.2 |
| DNMT3A | mutated | c.1517A>G | p.His506Arg | 2.6 |
| DNMT3A | mutated | c.1523T>C | p.Leu508Pro | 1.5 |
| DNMT3A | mutated | c.1543C>T | p.Gln515* | 19.4 |
| DNMT3A | mutated | c.1543del | p.Gln515Lysfs*136 | 1.8 |
| DNMT3A | mutated | c.1551_1552delinsG | p.Cys517Trpfs*134 | 1.3 |
| DNMT3A | mutated | c.1555-2A>T | p.splice site mutation | 4.8 |
| DNMT3A | mutated | c.1555-8_1555del | p.splice site mutation | 1.7 |
| DNMT3A | mutated | c.1591G>A | p.Asp531Asn | 1.4 |
| DNMT3A | mutated | c.1592A>G | p.Asp531Gly | 6.1 |
| DNMT3A | mutated | c.1628G>C | p.Gly543Ala | 21.6 |
| DNMT3A | mutated | c.1637T>A | p.Val546Glu | 3.8 |
| DNMT3A | mutated | c.1640T>A | p.Leu547His | 4.1 |
| DNMT3A | mutated | c.1640T>A | p.Leu547His | 1.4 |
| DNMT3A | mutated | c.1657_1659del | p.Asn553del | 8.9 |
| DNMT3A | mutated | c.1713_1724del | p.Ala572_Ala575del | 1.4 |
| DNMT3A | mutated | c.1726_1729delinsC | p.Ile576_Lys577delinsGln | 22.6 |
| DNMT3A | mutated | c.1742G>A | p.Trp581* | 1.8 |
| DNMT3A | mutated | c.1903C>T | p.Arg635Trp | 2.8 |

| | | | | |
|---|---|---|---|---|
| DNMT3A | mutated | c.1906G>A | p.Val636Met | 1.1 |
| DNMT3A | mutated | c.1969G>A | p.Val657Met | 5.3 |
| DNMT3A | mutated | c.1969G>A | p.Val657Met | 1.4 |
| DNMT3A | mutated | c.1972G>T | p.Asp658Tyr | 10.0 |
| DNMT3A | mutated | c.1976G>A | p.Arg659His | 1.4 |
| DNMT3A | mutated | c.1979A>G | p.Tyr660Cys | 4.5 |
| DNMT3A | mutated | c.1998_1999del | p.Cys666* | 2.2 |
| DNMT3A | mutated | c.1998T>G | p.Cys666Trp | 2.9 |
| DNMT3A | mutated | c.2007dup | p.Ile670Hisfs*43 | 20.6 |
| DNMT3A | mutated | c.2023G>A | p.Val675Met | 1.1 |
| DNMT3A | mutated | c.2024_2026dup | p.Val675_Arg676insLeu | 4.7 |
| DNMT3A | mutated | c.2032del | p.Gln678Argfs*27 | 1.1 |
| DNMT3A | mutated | c.2053G>C | p.Gly685Arg | 15.0 |
| DNMT3A | mutated | c.2056del | p.Asp686Thrfs*19 | 1.9 |
| DNMT3A | mutated | c.2057A>G | p.Asp686Gly | 2.3 |
| DNMT3A | mutated | c.2062C>T | p.Arg688Cys | 1.7 |
| DNMT3A | mutated | c.2063G>A | p.Arg688His | 1.1 |
| DNMT3A | mutated | c.2063G>A | p.Arg688His | 2.3 |
| DNMT3A | mutated | c.2084T>C | p.Ile695Thr | 2.0 |
| DNMT3A | mutated | c.2095G>C | p.Gly699Arg | 16.7 |
| DNMT3A | mutated | c.2095G>C | p.Gly699Arg | 2.3 |
| DNMT3A | mutated | c.2098C>A | p.Pro700Thr | 4.7 |
| DNMT3A | mutated | c.2099C>T | p.Pro700Leu | 2.4 |
| DNMT3A | mutated | c.2104G>T | p.Asp702Tyr | 11.8 |
| DNMT3A | mutated | c.2108del | p.Leu703Argfs*2 | 1.1 |
| DNMT3A | mutated | c.2114T>C | p.Ile705Thr | 1.3 |
| DNMT3A | mutated | c.2171A>G | p.Tyr724Cys | 2.2 |
| DNMT3A | mutated | c.2171A>G | p.Tyr724Cys | 5.7 |
| DNMT3A | mutated | c.2177G>T | p.Gly726Val | 1.7 |
| DNMT3A | mutated | c.2183G>A | p.Gly728Asp | 5.6 |
| DNMT3A | mutated | c.2185C>T | p.Arg729Trp | 30.8 |
| DNMT3A | mutated | c.2185C>T | p.Arg729Trp | 13.9 |
| DNMT3A | mutated | c.2192T>A | p.Phe731Tyr | 5.0 |
| DNMT3A | mutated | c.2204A>C | p.Tyr735Ser | 8.5 |
| DNMT3A | mutated | c.2204A>G | p.Tyr735Cys | 1.1 |
| DNMT3A | mutated | c.2204A>G | p.Tyr735Cys | 11.3 |
| DNMT3A | mutated | c.2204A>G | p.Tyr735Cys | 22.8 |
| DNMT3A | mutated | c.2204A>G | p.Tyr735Cys | 1.4 |
| DNMT3A | mutated | c.2204A>G | p.Tyr735Cys | 1.9 |
| DNMT3A | mutated | c.2206C>T | p.Arg736Cys | 9.6 |
| DNMT3A | mutated | c.2206C>T | p.Arg736Cys | 4.8 |
| DNMT3A | mutated | c.2206C>T | p.Arg736Cys | 9.5 |
| DNMT3A | mutated | c.2228C>T | p.Pro743Leu | 12.2 |
| DNMT3A | mutated | c.2245C>T | p.Arg749Cys | 5.6 |
| DNMT3A | mutated | c.2245C>T | p.Arg749Cys | 6.3 |
| DNMT3A | mutated | c.2245C>T | p.Arg749Cys | 4.4 |
| DNMT3A | mutated | c.2254T>G | p.Phe752Val | 1.0 |

| | | | | |
|---|---|---|---|---|
| DNMT3A | mutated | c.2259G>A | p.Trp753* | 5.2 |
| DNMT3A | mutated | c.2261T>C | p.Leu754Pro | 1.8 |
| DNMT3A | mutated | c.2261T>G | p.Leu754Arg | 1.8 |
| DNMT3A | mutated | c.2264T>C | p.Phe755Ser | 4.3 |
| DNMT3A | mutated | c.2302delG | p.Asp768Thrfs*11 | 2.3 |
| DNMT3A | mutated | c.2309C>T | p.Ser770Leu | 2.9 |
| DNMT3A | mutated | c.2311C>G | p.Arg771Gly | 3.6 |
| DNMT3A | mutated | c.2311C>T | p.Arg771* | 4.9 |
| DNMT3A | mutated | c.2311C>T | p.Arg771* | 15.3 |
| DNMT3A | mutated | c.2311C>T | p.Arg771* | 1.3 |
| DNMT3A | mutated | c.2311C>T | p.Arg771* | 1.8 |
| DNMT3A | mutated | c.2312G>T | p.Arg771Leu | 23.7 |
| DNMT3A | mutated | c.2320G>T | p.Glu774* | 11.3 |
| DNMT3A | mutated | c.2330C>G | p.Pro777Arg | 1.3 |
| DNMT3A | mutated | c.2332G>A | p.Val778Met | 1.3 |
| DNMT3A | mutated | c.2333T>G | p.Val778Gly | 6.5 |
| DNMT3A | mutated | c.2333T>G | p.Val778Gly | 2.8 |
| DNMT3A | mutated | c.2339T>C | p.Ile780Thr | 5.4 |
| DNMT3A | mutated | c.2339T>C | p.Ile780Thr | 1.1 |
| DNMT3A | mutated | c.2339T>C | p.Ile780Thr | 2.2 |
| DNMT3A | mutated | c.2347A>T | p.Lys783* | 1.6 |
| DNMT3A | mutated | c.2377T>G | p.Tyr793Asp | 1.3 |
| DNMT3A | mutated | c.2387del | p.Gly796Valfs*6 | 2.2 |
| DNMT3A | mutated | c.2387G>A | p.Gly796Asp | 1.5 |
| DNMT3A | mutated | c.2389A>T | p.Asn797Tyr | 2.7 |
| DNMT3A | mutated | c.2390A>G | p.Asn797Ser | 17.0 |
| DNMT3A | mutated | c.2395C>T | p.Pro799Ser | 5.0 |
| DNMT3A | mutated | c.2404A>T | p.Asn802Tyr | 5.1 |
| DNMT3A | mutated | c.2462del | p.His821Leufs*4 | 3.0 |
| DNMT3A | mutated | c.2471dup | p.Ala825Serfs*30 | 1.4 |
| DNMT3A | mutated | c.2524C>T | p.Gln842* | 2.4 |
| DNMT3A | mutated | c.2524C>T | p.Gln842* | 3.7 |
| DNMT3A | mutated | c.2533del | p.Asp845Thrfs*8 | 3.0 |
| DNMT3A | mutated | c.2550del | p.Phe851Serfs*2 | 3.1 |
| DNMT3A | mutated | c.2578T>C | p.Trp860Arg | 6.2 |
| DNMT3A | mutated | c.2617C>T | p.His873Tyr | 1.1 |
| DNMT3A | mutated | c.2644C>T | p.Arg882Cys | 2.6 |
| DNMT3A | mutated | c.2644C>T | p.Arg882Cys | 32.2 |
| DNMT3A | mutated | c.2645G>A | p.Arg882His | 2.2 |
| DNMT3A | mutated | c.2645G>A | p.Arg882His | 2.5 |
| DNMT3A | mutated | c.2645G>A | p.Arg882His | 3.2 |
| DNMT3A | mutated | c.2645G>A | p.Arg882His | 1.5 |
| DNMT3A | mutated | c.2666T>C | p.Leu889Pro | 2.2 |
| DNMT3A | mutated | c.2666T>C | p.Leu889Pro | 0.9 |
| DNMT3A | mutated | c.2679G>C | p.Trp893Cys | 3.5 |
| DNMT3A | mutated | c.2695C>T | p.Arg899Cys | 1.9 |
| DNMT3A | mutated | c.2695C>T | p.Arg899Cys | 2.0 |

| | | | | |
|---|---|---|---|---|
| DNMT3A | mutated | c.2695C>T | p.Arg899Cys | 4.7 |
| DNMT3A | mutated | c.2695C>T | p.Arg899Cys | 2.2 |
| DNMT3A | mutated | c.2705del | p.Phe902Serfs*4 | 8.2 |
| DNMT3A | mutated | c.2710C>T | p.Pro904Ser | 1.2 |
| DNMT3A | mutated | c.2714T>G | p.Leu905Arg | 1.6 |
| DNMT3A | mutated | c.2723A>G | p.Tyr908Cys | 1.9 |
| DNMT3A | mutated | c.2726T>C | p.Phe909Ser | 1.8 |
| DNMT3A | mutated | c.719_725del | p.Glu240Alafs*74 | 1.8 |
| DNMT3A | mutated | c.814A>T | p.Lys272* | 1.1 |
| DNMT3A | mutated | c.884T>A | p.Leu295Gln | 2.3 |
| DNMT3A | mutated | c.890G>A | p.Trp297* | 1.5 |
| DNMT3A | mutated | c.893G>A | p.Gly298Glu | 2.6 |
| DNMT3A | mutated | c.901C>T | p.Arg301Trp | 2.2 |
| DNMT3A | mutated | c.905G>T | p.Gly302Val | 13.9 |
| DNMT3A | mutated | c.914G>A | p.Trp305* | 2.5 |
| DNMT3A | mutated | c.915G>A | p.Trp305* | 26.4 |
| DNMT3A | mutated | c.929T>C | p.Ile310Thr | 3.0 |
| DNMT3A | mutated | c.976C>T | p.Arg326Cys | 20.5 |
| DNMT3A | mutated | c.976C>T | p.Arg326Cys | 3.6 |
| DNMT3A | mutated | c.976C>T | p.Arg326Cys | 8.4 |
| DNMT3A | mutated | c.98G>A | p.Arg33His | 1.8 |
| EZH2 | mutated | c.875A>G | p.Tyr292Cys | 1.1 |
| GNAS | mutated | c.2531G>A | p.Arg844His | 2.9 |
| GNB1 | mutated | c.169A>G | p.Lys57Glu | 13.9 |
| IDH1 | mutated | c.395G>A | p.Arg132His | 43.7 |
| JAK2 | mutated | c.1849G>T | p.Val617Phe | 2.0 |
| JAK2 | mutated | c.1849G>T | p.Val617Phe | 2.6 |
| JAK2 | mutated | c.1849G>T | p.Val617Phe | 5.5 |
| JAK2 | mutated | c.1849G>T | p.Val617Phe | 2.3 |
| JAK2 | mutated | c.1849G>T | p.Val617Phe | 4.5 |
| JAK2 | mutated | c.1849G>T | p.Val617Phe | 38.7 |
| JAK2 | mutated | c.1849G>T | p.Val617Phe | 1.5 |
| JAK2 | mutated | c.2569A>G | p.Lys857Glu | 1.2 |
| KRAS | mutated | c.35G>A | p.Gly12Asp | 4.3 |
| PPM1D | mutated | c.1270G>T | p.Glu424* | 5.9 |
| PPM1D | mutated | c.1297A>T | p.Lys433* | 1.0 |
| PPM1D | mutated | c.1349del | p.Leu450* | 4.4 |
| PPM1D | mutated | c.1372C>T | p.Arg458* | 17.4 |
| PPM1D | mutated | c.1382del | p.Val461Alafs*4 | 2.4 |
| PPM1D | mutated | c.1434C>A | p.Cys478* | 8.8 |
| PPM1D | mutated | c.1528C>T | p.Gln510* | 2.8 |
| PPM1D | mutated | c.1535del | p.Asn512Ilefs*2 | 0.7 |
| PPM1D | mutated | c.1535del | p.Asn512Ilefs*2 | 0.9 |
| PPM1D | mutated | c.1566dup | p.Ala523Serfs*5 | 10.0 |
| PPM1D | mutated | c.1609del | p.Thr537Hisfs*2 | 34.0 |
| PPM1D | mutated | c.1649del | p.His550Leufs*6 | 6.9 |
| PPM1D | mutated | c.1654C>T | p.Arg552* | 2.7 |

| | | | | |
|---|---|---|---|---|
| PPM1D | mutated | c.1654C>T | p.Arg552* | 13.6 |
| PPM1D | mutated | c.1714C>T | p.Arg572* | 2.8 |
| RAD21 | mutated | c.144+1G>T | p.splice site mutation | 6.1 |
| SETBP1 | mutated | c.2572G>A | p.Glu858Lys | 1.8 |
| SF3B1 | mutated | c.1868A>T | p.Tyr623Phe | 1.3 |
| SF3B1 | mutated | c.1876A>G | p.Asn626Asp | 1.3 |
| SF3B1 | mutated | c.1985A>T | p.His662Leu | 1.0 |
| SF3B1 | mutated | c.1986C>G | p.His662Gln | 1.2 |
| SF3B1 | mutated | c.1996A>C | p.Lys666Gln | 4.8 |
| SF3B1 | mutated | c.1997A>G | p.Lys666Arg | 7.8 |
| SF3B1 | mutated | c.1998G>C | p.Lys666Asn | 8.1 |
| SF3B1 | mutated | c.1998G>C | p.Lys666Asn | 1.0 |
| SF3B1 | mutated | c.1998G>T | p.Lys666Asn | 5.4 |
| SF3B1 | mutated | c.1998G>T | p.Lys666Asn | 27.1 |
| SF3B1 | mutated | c.1998G>T | p.Lys666Asn | 23.9 |
| SF3B1 | mutated | c.1998G>T | p.Lys666Asn | 1.7 |
| SF3B1 | mutated | c.1998G>T | p.Lys666Asn | 26.7 |
| SF3B1 | mutated | c.2098A>G | p.Lys700Glu | 13.8 |
| SF3B1 | mutated | c.2098A>G | p.Lys700Glu | 23.0 |
| SMC1A | mutated | c.2131C>T | p.Arg711Trp | 4.7 |
| SMC3 | mutated | c.3353G>T | p.Gly1118Val | 0.7 |
| SMC3 | mutated | c.3598G>A | p.Val1200Met | 6.0 |
| SRSF2 | mutated | c.170T>A | p.Phe57Tyr | 2.0 |
| SRSF2 | mutated | c.170T>A | p.Phe57Tyr | 2.1 |
| SRSF2 | mutated | c.284C>A | p.Pro95His | 32.0 |
| SRSF2 | mutated | c.284C>A | p.Pro95His | 23.1 |
| SRSF2 | mutated | c.284C>G | p.Pro95Arg | 1.2 |
| SRSF2 | mutated | c.284C>G | p.Pro95Arg | 18.2 |
| SRSF2 | mutated | c.284C>T | p.Pro95Leu | 41.8 |
| SRSF2 | mutated | c.287C>T | p.Pro96Leu | 7.4 |
| TET2 | mutated | c.1028_1046del | p.Thr343Metfs*23 | 2.4 |
| TET2 | mutated | c.1061C>A | p.Ser354* | 17.5 |
| TET2 | mutated | c.1061C>G | p.Ser354* | 1.3 |
| TET2 | mutated | c.1201_1203del | p.Pro401del | 2.7 |
| TET2 | mutated | c.1201_1203del | p.Pro401del | 2.8 |
| TET2 | mutated | c.1212del | p.Leu404Phefs*23 | 1.9 |
| TET2 | mutated | c.1219del | p.Ser407Leufs*20 | 27.7 |
| TET2 | mutated | c.1249C>T | p.Gln417* | 4.4 |
| TET2 | mutated | c.1259del | p.Ser420* | 1.6 |
| TET2 | mutated | c.1430del | p.Ser477Leufs*9 | 3.2 |
| TET2 | mutated | c.1469_1470del | p.Ile490Thrfs*13 | 1.7 |
| TET2 | mutated | c.1588C>T | p.Gln530* | 5.9 |
| TET2 | mutated | c.1630C>T | p.Arg544* | 25.0 |
| TET2 | mutated | c.1630C>T | p.Arg544* | 3.8 |
| TET2 | mutated | c.1699_1703del | p.Leu567Glyfs*14 | 2.7 |
| TET2 | mutated | c.1800_1801dup | p.Thr601Argfs*39 | 6.3 |
| TET2 | mutated | c.1803del | p.Ser602Profs*37 | 16.5 |

| | | | | |
|---|---|---|---|---|
| TET2 | mutated | c.1842del | p.Leu615Serfs*24 | 3.0 |
| TET2 | mutated | c.1863_1879del | p.Gln622Glyfs*10 | 3.8 |
| TET2 | mutated | c.2167_2170del | p.Pro723Ilefs*27 | 1.8 |
| TET2 | mutated | c.2255_2261del | p.Asn752Argfs*59 | 2.1 |
| TET2 | mutated | c.2276del | p.Thr759Ilefs*54 | 5.5 |
| TET2 | mutated | c.2370_2382dup | p.Ser795Valfs*11 | 6.6 |
| TET2 | mutated | c.2375C>G | p.Ser792* | 17.4 |
| TET2 | mutated | c.2662C>T | p.Gln888* | 2.6 |
| TET2 | mutated | c.2662C>T | p.Gln888* | 6.0 |
| TET2 | mutated | c.2674C>T | p.Gln892* | 7.6 |
| TET2 | mutated | c.2746C>T | p.Gln916* | 4.6 |
| TET2 | mutated | c.2746C>T | p.Gln916* | 5.1 |
| TET2 | mutated | c.2746C>T | p.Gln916* | 1.5 |
| TET2 | mutated | c.2749C>T | p.Gln917* | 27.0 |
| TET2 | mutated | c.2757C>A | p.Tyr919* | 1.8 |
| TET2 | mutated | c.2839C>T | p.Gln947* | 19.2 |
| TET2 | mutated | c.2884C>T | p.Gln962* | 29.5 |
| TET2 | mutated | c.2896C>T | p.Gln966* | 2.0 |
| TET2 | mutated | c.2905C>T | p.Gln969* | 1.3 |
| TET2 | mutated | c.2926C>T | p.Gln976* | 2.0 |
| TET2 | mutated | c.2926C>T | p.Gln976* | 16.3 |
| TET2 | mutated | c.2944A>T | p.Lys982* | 38.9 |
| TET2 | mutated | c.3119T>G | p.Leu1040* | 2.9 |
| TET2 | mutated | c.3127del | p.His1043Ilefs*12 | 21.0 |
| TET2 | mutated | c.3287del | p.Thr1096Lysfs*10 | 2.6 |
| TET2 | mutated | c.3339_3340del | p.Thr1114Serfs*15 | 1.3 |
| TET2 | mutated | c.3344del | p.Pro1115Leufs*2 | 1.9 |
| TET2 | mutated | c.3344del | p.Pro1115Leufs*2 | 1.0 |
| TET2 | mutated | c.3369del | p.Val1124Serfs*13 | 2.0 |
| TET2 | mutated | c.3404G>A | p.Cys1135Tyr | 1.9 |
| TET2 | mutated | c.3404G>A | p.Cys1135Tyr | 1.8 |
| TET2 | mutated | c.3409+1G>A | p.splice site mutation | 2.6 |
| TET2 | mutated | c.3415del | p.Ile1139Leufs*13 | 6.9 |
| TET2 | mutated | c.3415del | p.Ile1139Leufs*13 | 3.1 |
| TET2 | mutated | c.3491T>G | p.Met1164Arg | 1.0 |
| TET2 | mutated | c.3500G>A | p.Arg1167Lys | 1.1 |
| TET2 | mutated | c.3522_3523insG | p.Ile1175Aspfs*2 | 19.0 |
| TET2 | mutated | c.3523A>T | p.Ile1175Phe | 18.7 |
| TET2 | mutated | c.3524_3526delinsCTT | p.Ile1175_Arg1176delinsThrTrp | 5.9 |
| TET2 | mutated | c.3530T>G | p.Ile1177Ser | 7.4 |
| TET2 | mutated | c.3578G>A | p.Cys1193Tyr | 2.6 |
| TET2 | mutated | c.3637G>A | p.Val1213Met | 1.4 |
| TET2 | mutated | c.3640C>T | p.Arg1214Trp | 1.4 |
| TET2 | mutated | c.3640C>T | p.Arg1214Trp | 4.7 |
| TET2 | mutated | c.3646C>T | p.Arg1216* | 1.5 |
| TET2 | mutated | c.3656A>C | p.His1219Pro | 1.4 |
| TET2 | mutated | c.3661T>G | p.Cys1221Gly | 1.7 |

| | | | | |
|---|---|---|---|---|
| TET2 | mutated | c.3662G>C | p.Cys1221Ser | 1.5 |
| TET2 | mutated | c.3732_3733del | p.Tyr1245Leufs*22 | 4.8 |
| TET2 | mutated | c.3732_3733del | p.Tyr1245Leufs*22 | 1.5 |
| TET2 | mutated | c.3733_3737del | p.Tyr1245Glyfs*21 | 1.2 |
| TET2 | mutated | c.3734A>G | p.Tyr1245Cys | 39.6 |
| TET2 | mutated | c.3755T>C | p.Leu1252Pro | 4.3 |
| TET2 | mutated | c.3781C>T | p.Arg1261Cys | 1.6 |
| TET2 | mutated | c.3782G>A | p.Arg1261His | 2.0 |
| TET2 | mutated | c.3785G>A | p.Arg1262Gln | 2.0 |
| TET2 | mutated | c.3788G>C | p.Cys1263Ser | 1.5 |
| TET2 | mutated | c.3788G>C | p.Cys1263Ser | 1.6 |
| TET2 | mutated | c.3821_3822del | p.Gln1274Argfs*25 | 1.5 |
| TET2 | mutated | c.3822G>C | p.Gln1274His | 5.7 |
| TET2 | mutated | c.3863G>A | p.Gly1288Asp | 2.0 |
| TET2 | mutated | c.3863G>A | p.Gly1288Asp | 3.3 |
| TET2 | mutated | c.3866G>T | p.Cys1289Phe | 2.2 |
| TET2 | mutated | c.3894dup | p.Lys1299* | 13.5 |
| TET2 | mutated | c.3904A>G | p.Arg1302Gly | 5.7 |
| TET2 | mutated | c.3968del | p.Glu1323Glyfs*40 | 37.8 |
| TET2 | mutated | c.4015A>T | p.Lys1339* | 23.3 |
| TET2 | mutated | c.4021dup | p.Ala1341Glyfs*3 | 1.6 |
| TET2 | mutated | c.4030G>A | p.Ala1344Thr | 8.0 |
| TET2 | mutated | c.4042del | p.Gln1348Argfs*15 | 7.6 |
| TET2 | mutated | c.4075C>A | p.Arg1359Ser | 4.7 |
| TET2 | mutated | c.4075C>A | p.Arg1359Ser | 1.2 |
| TET2 | mutated | c.4076G>A | p.Arg1359His | 2.9 |
| TET2 | mutated | c.4081G>C | p.Gly1361Arg | 4.9 |
| TET2 | mutated | c.4082G>A | p.Gly1361Asp | 2.9 |
| TET2 | mutated | c.4103_4116del | p.Phe1368Cysfs*28 | 19.2 |
| TET2 | mutated | c.4126G>A | p.Asp1376Asn | 1.9 |
| TET2 | mutated | c.4131_4132del | p.Phe1377Leufs*23 | 21.5 |
| TET2 | mutated | c.4132T>C | p.Cys1378Arg | 19.6 |
| TET2 | mutated | c.4133G>A | p.Cys1378Tyr | 26.3 |
| TET2 | mutated | c.4136C>T | p.Ala1379Val | 36.6 |
| TET2 | mutated | c.4138C>T | p.His1380Tyr | 7.6 |
| TET2 | mutated | c.4138C>T | p.His1380Tyr | 1.5 |
| TET2 | mutated | c.4140T>G | p.His1380Gln | 5.9 |
| TET2 | mutated | c.4193T>G | p.Leu1398Arg | 6.4 |
| TET2 | mutated | c.4234G>T | p.Asp1412Tyr | 19.3 |
| TET2 | mutated | c.4256C>G | p.Pro1419Arg | 7.7 |
| TET2 | mutated | c.4354C>T | p.Arg1452* | 5.0 |
| TET2 | mutated | c.4393C>T | p.Arg1465* | 2.1 |
| TET2 | mutated | c.4393C>T | p.Arg1465* | 26.9 |
| TET2 | mutated | c.4399del | p.Arg1467Glyfs*3 | 14.6 |
| TET2 | mutated | c.4481C>G | p.Ser1494* | 3.6 |
| TET2 | mutated | c.4546C>T | p.Arg1516* | 29.7 |
| TET2 | mutated | c.4546C>T | p.Arg1516* | 3.9 |

| | | | | |
|---|---|---|---|---|
| TET2 | mutated | c.4570C>T | p.Gln1524* | 12.3 |
| TET2 | mutated | c.4589_4618del | p.Pro1530_Gln1539del | 57.0 |
| TET2 | mutated | c.4621C>T | p.Gln1541* | 18.4 |
| TET2 | mutated | c.4624C>T | p.Gln1542* | 6.6 |
| TET2 | mutated | c.4639C>T | p.Gln1547* | 1.8 |
| TET2 | mutated | c.4757C>G | p.Ser1586* | 3.0 |
| TET2 | mutated | c.4854C>G | p.Tyr1618* | 5.0 |
| TET2 | mutated | c.4879C>T | p.Gln1627* | 2.5 |
| TET2 | mutated | c.506_508delinsC | p.His169Profs*6 | 2.4 |
| TET2 | mutated | c.5220dup | p.Pro1741Thrfs*12 | 2.9 |
| TET2 | mutated | c.5271_5272dup | p.Ser1758Phefs*6 | 28.7 |
| TET2 | mutated | c.532G>T | p.Glu178* | 29.3 |
| TET2 | mutated | c.5413_5420del | p.Asn1805* | 3.7 |
| TET2 | mutated | c.5454_5458del | p.Leu1819* | 9.8 |
| TET2 | mutated | c.5467_5472delinsCC | p.Asn1823Profs*9 | 25.7 |
| TET2 | mutated | c.5482C>T | p.Gln1828* | 2.1 |
| TET2 | mutated | c.5500C>T | p.Gln1834* | 4.2 |
| TET2 | mutated | c.5541G>A | p.Trp1847* | 3.1 |
| TET2 | mutated | c.5541G>A | p.Trp1847* | 2.1 |
| TET2 | mutated | c.5543C>A | p.Ser1848* | 4.2 |
| TET2 | mutated | c.5551_5554del | p.Glu1851Argfs*35 | 4.0 |
| TET2 | mutated | c.5551G>T | p.Glu1851* | 0.8 |
| TET2 | mutated | c.5603A>G | p.His1868Arg | 2.1 |
| TET2 | mutated | c.5615T>A | p.Leu1872His | 3.3 |
| TET2 | mutated | c.561dup | p.Lys188Glufs*4 | 18.9 |
| TET2 | mutated | c.5621A>T | p.Glu1874Val | 2.1 |
| TET2 | mutated | c.5636A>C | p.Glu1879Ala | 11.5 |
| TET2 | mutated | c.5642A>G | p.His1881Arg | 2.5 |
| TET2 | mutated | c.5650A>G | p.Thr1884Ala | 1.4 |
| TET2 | mutated | c.5690T>G | p.Ile1897Ser | 4.1 |
| TET2 | mutated | c.5720T>A | p.Met1907Lys | 11.1 |
| TET2 | mutated | c.651del | p.Val218Trpfs*32 | 1.4 |
| TET2 | mutated | c.661_667del | p.Thr221Valfs*27 | 5.8 |
| TET2 | mutated | c.662_663insTC | p.Gly223Metfs*28 | 2.2 |
| TET2 | mutated | c.763C>T | p.Gln255* | 6.5 |
| TET2 | mutated | c.822del | p.Asn275Ilefs*18 | 2.0 |
| TET2 | mutated | c.822del | p.Asn275Ilefs*18 | 1.3 |
| TET2 | mutated | c.840_841insTT | p.Asn281Leufs*13 | 4.1 |
| TET2 | mutated | c.897dup | p.Asp300* | 4.9 |
| TP53 | mutated | c.223C>G | p.Pro75Ala | 4.7 |
| TP53 | mutated | c.329G>C | p.Arg110Pro | 5.2 |
| TP53 | mutated | c.332T>G | p.Leu111Arg | 1.7 |
| TP53 | mutated | c.376T>C | p.Tyr126His | 5.9 |
| TP53 | mutated | c.464C>T | p.Thr155Ile | 3.2 |
| TP53 | mutated | c.473G>T | p.Arg158Leu | 4.4 |
| TP53 | mutated | c.530C>T | p.Pro177Leu | 2.5 |
| TP53 | mutated | c.533A>C | p.His178Pro | 2.9 |

| | | | | |
|---|---|---|---|---|
| TP53 | mutated | c.541C>T | p.Arg181Cys | 45.9 |
| TP53 | mutated | c.584T>C | p.Ile195Thr | 11.4 |
| TP53 | mutated | c.586C>T | p.Arg196* | 24.2 |
| TP53 | mutated | c.658T>C | p.Tyr220His | 2.4 |
| TP53 | mutated | c.668C>T | p.Pro223Leu | 2.1 |
| TP53 | mutated | c.731G>T | p.Gly244Val | 1.9 |
| TP53 | mutated | c.734G>A | p.Gly245Asp | 4.6 |
| TP53 | mutated | c.745A>G | p.Arg249Gly | 2.6 |
| TP53 | mutated | c.817C>T | p.Arg273Cys | 1.7 |
| TP53 | mutated | c.997del | p.Arg333Valfs*12 | 2.5 |
| U2AF1 | mutated | c.101C>T | p.Ser34Phe | 7.0 |
| U2AF1 | mutated | c.470A>C | p.Gln157Pro | 9.8 |
| U2AF1 | mutated | c.470A>C | p.Gln157Pro | 3.2 |
| U2AF1 | mutated | c.470A>G | p.Gln157Arg | 1.6 |
| U2AF1 | mutated | c.470A>G | p.Gln157Arg | 23.6 |
| U2AF1 | mutated | c.470A>G | p.Gln157Arg | 4.6 |
| U2AF1 | mutated | c.470A>G | p.Gln157Arg | 37.1 |
| ZRSR2 | mutated | c.1017del | p.Trp340Glyfs*? | 20.9 |
| ZRSR2 | mutated | c.1141dup | p.Arg381Lysfs*4 | 16.9 |
| ZRSR2 | mutated | c.1223dup | p.His408Glnfs*20 | 5.7 |
| ZRSR2 | mutated | c.195_198del | p.Glu67Glyfs*10 | 1.8 |
| ZRSR2 | mutated | c.376C>T | p.Arg126* | 17.5 |
| ZRSR2 | mutated | c.398_399del | p.Glu133Glyfs*11 | 8.3 |
| ZRSR2 | mutated | c.593del | p.Pro198Leufs*40 | 4.5 |
| ZRSR2 | mutated | c.706T>A | p.Phe236Ile | 1.8 |
| ZRSR2 | mutated | c.80G>T | p.Arg27Leu | 6.8 |
| ZRSR2 | mutated | c.83dup | p.Lys29Glufs*26 | 4.6 |
| ZRSR2 | mutated | c.860_864delinsAAT | p.Phe287* | 2.4 |
| ZRSR2 | mutated | c.988C>T | p.His330Tyr | 1.6 |

**Supplemental Table 2** – List of 159 unique CHIP mutations identified in STARNET in the genes ASXL1, DNMT3A, JAK2 and TET2 based on whole-genome-sequencing with limited depth (35-fold). Provided are gene name, confirmation of CHIP mutation – polymorphisms, variants, synonymous and uncertain mutations were excluded, change on DNA level, change on amino acid (AA) level and variant allele frequency (VAF).

| Gene | CHIP result | DNA result | AA result | VAF (%) |
|---|---|---|---|---|
| ASXL1 | mutated | c.1211C>T | p.Arg404* | 29.5 |
| ASXL1 | mutated | c.1550C>T | p.Gln517* | 20.8 |
| ASXL1 | mutated | c.2238C>T | p.Gln575* | 17.0 |
| ASXL1 | mutated | c.2331C>T | p.Arg606Trp | 43.9 |
| ASXL1 | mutated | c.2364A>G | p.Ile617Val | 42.9 |
| ASXL1 | mutated | c.2443G>T | p.Gly643Val | 67.6 |
| ASXL1 | mutated | c.2469G>A | p.Gly652Ser | 38.9 |
| ASXL1 | mutated | c.2469G>A | p.Gly652Ser | 46.7 |
| ASXL1 | mutated | c.2469G>A | p.Gly652Ser | 45.0 |
| ASXL1 | mutated | c.2469G>A | p.Gly652Ser | 53.3 |
| ASXL1 | mutated | c.2469G>A | p.Gly652Ser | 42.0 |
| ASXL1 | mutated | c.2469G>A | p.Gly652Ser | 53.3 |
| ASXL1 | mutated | c.2469G>A | p.Gly652Ser | 45.7 |
| ASXL1 | mutated | c.2469G>A | p.Gly652Ser | 59.3 |
| ASXL1 | mutated | c.2469G>A | p.Gly652Ser | 51.2 |
| ASXL1 | mutated | c.2469G>A | p.Gly652Ser | 68.8 |
| ASXL1 | mutated | c.2469G>A | p.Gly652Ser | 55.9 |
| ASXL1 | mutated | c.3223C>A | p.Ser903* | 15.0 |
| ASXL1 | mutated | c.3598C>T | p.Ser1028Leu | 72.7 |
| ASXL1 | mutated | c.3650G>C | p.Lys1045Asn | 61.2 |
| ASXL1 | mutated | c.3732C>T | p.Arg1073Cys | 28.8 |
| ASXL1 | mutated | c.4207C>T | p.Ser1231Phe | 57.4 |
| ASXL1 | mutated | c.4260A>G | p.Met1249Val | 31.5 |
| ASXL1 | mutated | c.4260A>G | p.Met1249Val | 50.0 |
| ASXL1 | mutated | c.4260A>G | p.Met1249Val | 43.2 |
| ASXL1 | mutated | c.4260A>G | p.Met1249Val | 60.6 |
| ASXL1 | mutated | c.4260A>G | p.Met1249Val | 48.1 |
| ASXL1 | mutated | c.4260A>G | p.Met1249Val | 56.7 |
| ASXL1 | mutated | c.4260A>G | p.Met1249Val | 52.7 |
| ASXL1 | mutated | c.4260A>G | p.Met1249Val | 53.2 |
| ASXL1 | mutated | c.4260A>G | p.Met1249Val | 47.4 |
| ASXL1 | mutated | c.4260A>G | p.Met1249Val | 47.5 |
| ASXL1 | mutated | c.4260A>G | p.Met1249Val | 55.4 |
| ASXL1 | mutated | c.4260A>G | p.Met1249Val | 60.0 |
| ASXL1 | mutated | c.4260A>G | p.Met1249Val | 39.0 |
| ASXL1 | mutated | c.4260A>G | p.Met1249Val | 53.1 |

| | | | | |
|---|---|---|---|---|
| ASXL1 | mutated | c.4260A>G | p.Met1249Val | 50.0 |
| ASXL1 | mutated | c.4260A>G | p.Met1249Val | 63.3 |
| ASXL1 | mutated | c.4630C>G | p.Thr1372Ser | 60.8 |
| ASXL1 | mutated | c.4630C>G | p.Thr1372Ser | 41.2 |
| ASXL1 | mutated | c.4630C>G | p.Thr1372Ser | 46.3 |
| ASXL1 | mutated | c.4698C>G | p.Leu1395Val | 40.4 |
| ASXL1 | mutated | c.4698C>G | p.Leu1395Val | 45.7 |
| ASXL1 | mutated | c.4704G>A | p.Gly1397Ser | 50.8 |
| ASXL1 | mutated | c.4890A>G | p.Ser1459Gly | 47.5 |
| ASXL1 | mutated | c.4894C>T | p.Ser1460Phe | 48.8 |
| ASXL1 | mutated | c.4980T>C | p.Ser1489Pro | 35.6 |
| ASXL1 | mutated | c.5137G>A | p.Arg1541Lys | 40.5 |
| ASXL1 | mutated | c.9387T>C | p.Val295Ala | 48.5 |
| DNMT3A | mutated | c.0498G>A | p.Arg326Cys | 20.0 |
| DNMT3A | mutated | c.0516G>A | p.Arg320* | 36.5 |
| DNMT3A | mutated | c.0555G>A | p.Pro307Ser | 34.1 |
| DNMT3A | mutated | c.0597C>A | p.Gly293Trp | 60.0 |
| DNMT3A | mutated | c.2050G>C | p.Ser786* | 50.0 |
| DNMT3A | mutated | c.2050G>A | p.Ser786Leu | 50.0 |
| DNMT3A | mutated | c.3054G>A | p.Thr44Met | 55.8 |
| DNMT3A | mutated | c.3568A>G | p.Ile705Thr | 16.7 |
| DNMT3A | mutated | c.4543A>G | p.Val657Ala | 25.5 |
| DNMT3A | mutated | c.7140C>T | p.Asp579Asn | 15.6 |
| DNMT3A | mutated | c.7290C>T | p.splice site mutation | 20.5 |
| DNMT3A | mutated | c.7290C>G | p.splice site mutation | 20.5 |
| DNMT3A | mutated | c.7476G>A | p.Gln534* | 36.4 |
| DNMT3A | mutated | c.8382G>C | p.Ser160Cys | 46.6 |
| DNMT3A | mutated | c.8648T>C | p.Gln842Arg | 44.4 |
| DNMT3A | mutated | c.9073G>A | p.Ala462Val | 43.5 |
| DNMT3A | mutated | c.9073G>A | p.Ala462Val | 62.7 |
| DNMT3A | mutated | c.9098C>A | p.Ala454Ser | 14.8 |
| DNMT3A | mutated | c.9529insC | p.Gly413fs* | 22.9 |
| DNMT3A | mutated | c.9529insC | p.Gly413fs* | 13.1 |
| DNMT3A | mutated | c.9529delG* | p.Gly413fs* | 12.0 |
| JAK2 | mutated | c.0232T>C | p.Leu712Pro | 46.5 |
| JAK2 | mutated | c.1803A>G | p.Asp838Gly | 47.7 |
| JAK2 | mutated | c.1828G>C | p.Glu846Asp | 50.0 |
| JAK2 | mutated | c.1828G>C | p.Glu846Asp | 53.8 |
| JAK2 | mutated | c.1828G>C | p.Glu846Asp | 46.5 |
| JAK2 | mutated | c.1828G>C | p.Glu846Asp | 61.4 |
| JAK2 | mutated | c.1828G>C | p.Glu846Asp | 48.6 |
| JAK2 | mutated | c.1828G>C | p.Glu846Asp | 48.6 |
| JAK2 | mutated | c.1828G>C | p.Glu846Asp | 50.9 |

| | | | | |
|---|---|---|---|---|
| JAK2 | mutated | c.2130G>A | p.Gly48Leu | 54.2 |
| JAK2 | mutated | c.2561G>A | p.Gly571Ser | 66.7 |
| JAK2 | mutated | c.2561G>A | p.Gly571Ser | 41.5 |
| JAK2 | mutated | c.2561G>A | p.Gly571Ser | 47.7 |
| JAK2 | mutated | c.2576A>G | p.Thr576Ala | 43.8 |
| JAK2 | mutated | c.2576A>G | p.Thr576Ala | 68.8 |
| JAK2 | mutated | c.3770G>T | p.Val617Phe | 32.6 |
| JAK2 | mutated | c.3770G>T | p.Val617Phe | 12.5 |
| JAK2 | mutated | c.3770G>T | p.Val617Phe | 34.1 |
| JAK2 | mutated | c.4997G>C | p.Ala391Pro | 56.4 |
| JAK2 | mutated | c.6715A>G | p.Asn1108Ser | 51.6 |
| JAK2 | mutated | c.6715A>G | p.Asn1108Ser | 55.3 |
| JAK2 | mutated | c.6715A>G | p.Asn1108Ser | 54.5 |
| JAK2 | mutated | c.6776C>G | p.Thr438Ser | 57.1 |
| JAK2 | mutated | c.7524A>C | p.Asn646His | 44.1 |
| JAK2 | mutated | c.7569A>G | p.Met661Val | 51.5 |
| JAK2 | mutated | c.9976C>G | p.Thr522Ser | 63.3 |
| TET2 | mutated | c.0785C>A | p.Cys1271* | 30.8 |
| TET2 | mutated | c.0798G>A | p.Arg1359His | 36.5 |
| TET2 | mutated | c.0843G>T | p.Cys1374Phe | 21.6 |
| TET2 | mutated | c.0852T>C | p.Tyr1294His | 50.0 |
| TET2 | mutated | c.4880G>A | p.Glu1250Lys | 64.5 |
| TET2 | mutated | c.4897C>G | p.Tyr1255* | 16.3 |
| TET2 | mutated | c.5620C>A | p.Pro14His | 46.7 |
| TET2 | mutated | c.5620C>A | p.Pro14His | 61.5 |
| TET2 | mutated | c.5620C>A | p.Pro14His | 48.8 |
| TET2 | mutated | c.5620C>A | p.Pro14His | 54.7 |
| TET2 | mutated | c.5620C>A | p.Pro14His | 59.6 |
| TET2 | mutated | c.5620C>A | p.Pro14His | 43.2 |
| TET2 | mutated | c.5620C>A | p.Pro14His | 47.6 |
| TET2 | mutated | c.5620C>A | p.Pro14His | 47.4 |
| TET2 | mutated | c.5620C>A | p.Pro14His | 54.2 |
| TET2 | mutated | c.5620C>A | p.Pro14His | 42.9 |
| TET2 | mutated | c.5620C>A | p.Pro14His | 45.5 |
| TET2 | mutated | c.5620C>A | p.Pro14His | 45.9 |
| TET2 | mutated | c.5620C>A | p.Pro14His | 64.3 |
| TET2 | mutated | c.5620C>A | p.Pro14His | 81.1 |
| TET2 | mutated | c.5620C>A | p.Pro14His | 51.8 |
| TET2 | mutated | c.5620C>A | p.Pro14His | 53.1 |
| TET2 | mutated | c.5800A>G | p.Tyr234Cys | 49.0 |
| TET2 | mutated | c.5843C>A | p.His248Gln | 38.6 |
| TET2 | mutated | c.5843C>A | p.His248Gln | 57.5 |
| TET2 | mutated | c.5890T>A | p.Ile264Asn | 51.7 |

| | | | | |
|---|---|---|---|---|
| TET2 | mutated | c.6209T>G | p.Tyr370* | 35.9 |
| TET2 | mutated | c.6213C>T | p.Arg1516* | 19.6 |
| TET2 | mutated | c.6384G>A | p.Gly429Arg | 47.2 |
| TET2 | mutated | c.6384G>A | p.Gly429Arg | 57.5 |
| TET2 | mutated | c.6770G>A | p.Met1701Ile | 50.0 |
| TET2 | mutated | c.6770G>A | p.Met1701Ile | 64.9 |
| TET2 | mutated | c.6770G>A | p.Met1701Ile | 50.0 |
| TET2 | mutated | c.6770G>A | p.Met1701Ile | 45.5 |
| TET2 | mutated | c.6770G>A | p.Met1701Ile | 58.1 |
| TET2 | mutated | c.6770G>A | p.Met1701Ile | 40.0 |
| TET2 | mutated | c.6770G>A | p.Met1701Ile | 44.7 |
| TET2 | mutated | c.6770G>A | p.Met1701Ile | 42.5 |
| TET2 | mutated | c.6770G>A | p.Met1701Ile | 60.5 |
| TET2 | mutated | c.6819C>G | p.Gln574Glu | 28.3 |
| TET2 | mutated | c.6819G>T | p.Val1718Leu | 56.4 |
| TET2 | mutated | c.6819G>T | p.Val1718Leu | 56.0 |
| TET2 | mutated | c.6819G>T | p.Val1718Leu | 51.2 |
| TET2 | mutated | c.6834C>T | p.Pro1723Ser | 44.9 |
| TET2 | mutated | c.6834C>T | p.Pro1723Ser | 52.6 |
| TET2 | mutated | c.6834C>T | p.Pro1723Ser | 64.3 |
| TET2 | mutated | c.6834C>T | p.Pro1723Ser | 48.8 |
| TET2 | mutated | c.6834C>T | p.Pro1723Ser | 48.6 |
| TET2 | mutated | c.6834C>T | p.Pro1723Ser | 60.4 |
| TET2 | mutated | c.7243G>GGTAA | p.splice site mutation | 43.8 |
| TET2 | mutated | c.7434G>T | p.Glu1923* | 51.4 |
| TET2 | mutated | c.7600T>C | p.Val1978Ala | 59.1 |
| TET2 | mutated | c.7600T>C | p.Val1978Ala | 46.5 |
| TET2 | mutated | c.7698T>C | p.Tyr867His | 59.1 |
| TET2 | mutated | c.7698T>C | p.Tyr867His | 52.9 |
| TET2 | mutated | c.7698T>C | p.Tyr867His | 42.5 |
| TET2 | mutated | c.7698T>C | p.Tyr867His | 50.0 |
| TET2 | mutated | c.7698T>C | p.Tyr867His | 57.4 |
| TET2 | mutated | c.7698T>C | p.Tyr867His | 50.0 |
| TET2 | mutated | c.7703T>G | p.Phe868Leu | 43.2 |
| TET2 | mutated | c.7785C>A | p.Leu896Ile | 55.7 |
| TET2 | mutated | c.8332C>T | p.Thr1078Ile | 49.1 |
| TET2 | mutated | c.8350A>C | p.Gln1084Pro | 56.3 |

**Supplemental Table 3 –** Patient characteristics of TET2 mutation carriers with CHIP-affected macrophages and controls without CHIP mutation based on STARNET. Matching between cases and controls was based on age and sex. Cardiovascular-relevant phenotype data are indicated. *: p<0.05.

| Baseline characteristics | TET2 CHIP (n=3) | TET2 non-CHIP (=21) | p-value |
|---|---|---|---|
| Age, mean (sd) | 60.3 (12.0) | 61.8 (8.2) | 0.82 |
| Sex (male) | 3 | 21 | 1 |
| BMI, mean (sd) | 28.0 (4.9) | 28.6 (5.0) | 0.86 |
| Arterial hypertension | 2 | 14 (n=20) | 1 |
| Hypercholesterolemia | 0 | 14 (n=20) | <0.05* |
| Smoking (ever) | 2 | 7 (n=20) | 0.54 |
| Diabetes | 0 | 4 (n=20) | 1 |
| Prior myocardial infarction | 1 | 7 (n=20) | 1 |
| Prior stroke | 0 | 0 (n=20) | 1 |

**Supplemental Table 4 –** Patient characteristics of ASXL1 mutation carriers with CHIP-affected macrophages and controls without CHIP mutation based on STARNET. Matching between cases and controls was based on age and sex. Cardiovascular-relevant phenotype data are indicated. *: p<0.05.

| **Baseline characteristics** | **ASXL1 CHIP (n=3)** | **ASXL1 non-CHIP (n=27)** | **p-value** |
|---|---|---|---|
| Age, mean (sd) | 67.3 (5.5) | 67.3 (5.6) | 0.98 |
| Sex (male) | 3 | 27 | 1 |
| BMI, mean (sd) | 26.4 (0.9) | 29.3 (3.6) | <0.05* |
| Arterial hypertension | 1 | 23 (n=26) | 0.07 |
| Hypercholesterolemia | 2 | 14 (n=26) | 1 |
| Smoking (ever) | 1 | 4 (n=26) | 0.45 |
| Diabetes | 0 | 4 (n=26) | 1 |
| Prior myocardial infarction | 1 | 10 (n=26) | 1 |
| Prior stroke | 0 | 4 (n=26) | 1 |

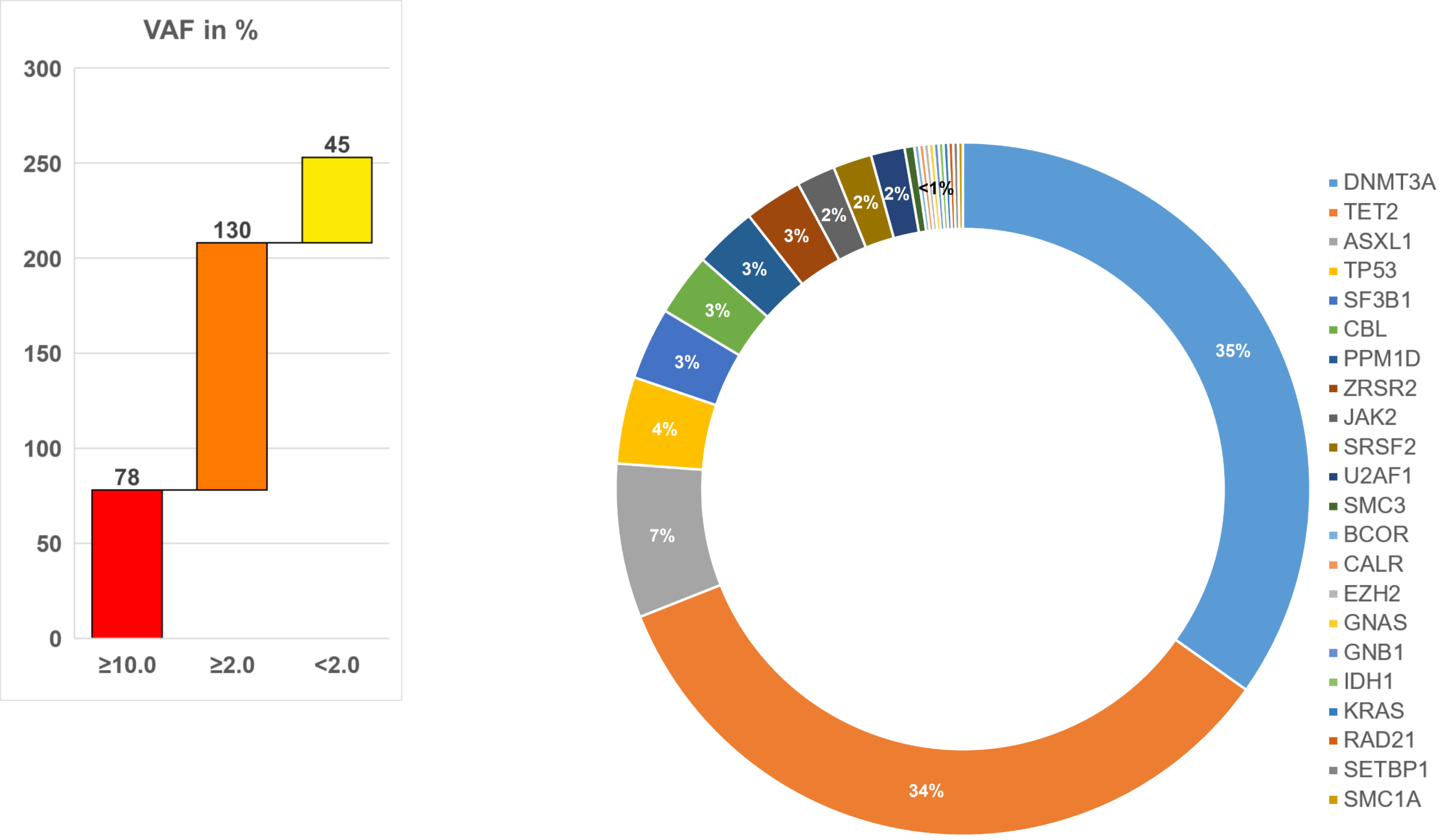


**Supplemental Figure 1** – VAF and distribution of CHIP mutations in MISSION. The left panel shows the distribution of VAF in 253 CHIP mutation carriers. For individuals with multiple CHIP mutations, the mutation with the highest VAF was considered. The right panel shows the percentage of genes affected by CHIP mutations. All 445 mutations identified by the 56-gene or 13-gene panel (Material and Methods Section) were considered. The color coding of the individual genes is indicated on the right.

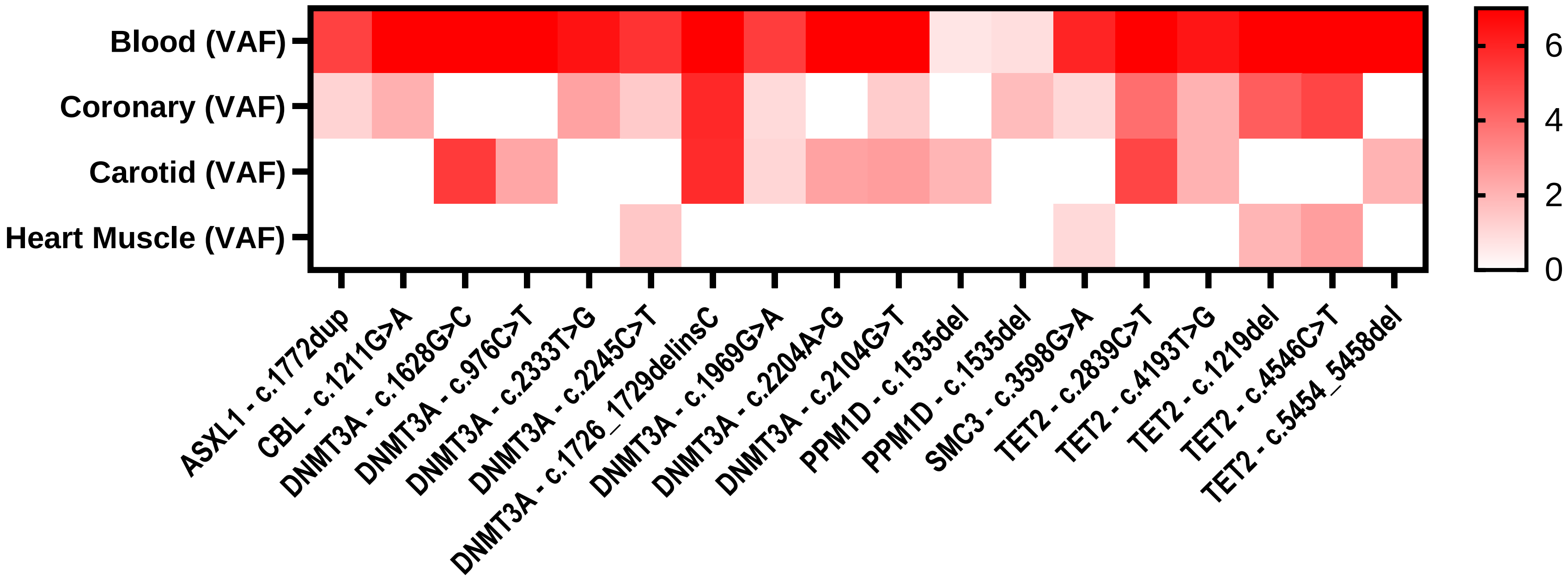


**Supplemental Figure 2** – DeepDNAseq identifies CHIP mutations in atherosclerotic coronary and carotid samples, and left ventricular myocardium. To confirm the identified CHIP mutations from whole blood in cardiovascular relevant tissues at the DNA level, coronary artery and carotid artery affected by atherosclerosis as well as myocardium of the left ventricle of CHIP mutation carriers were examined. The representation is in the format of a heat map. VAF >6 is presented in dark red. Tissues without mutation evidence are presented in white.

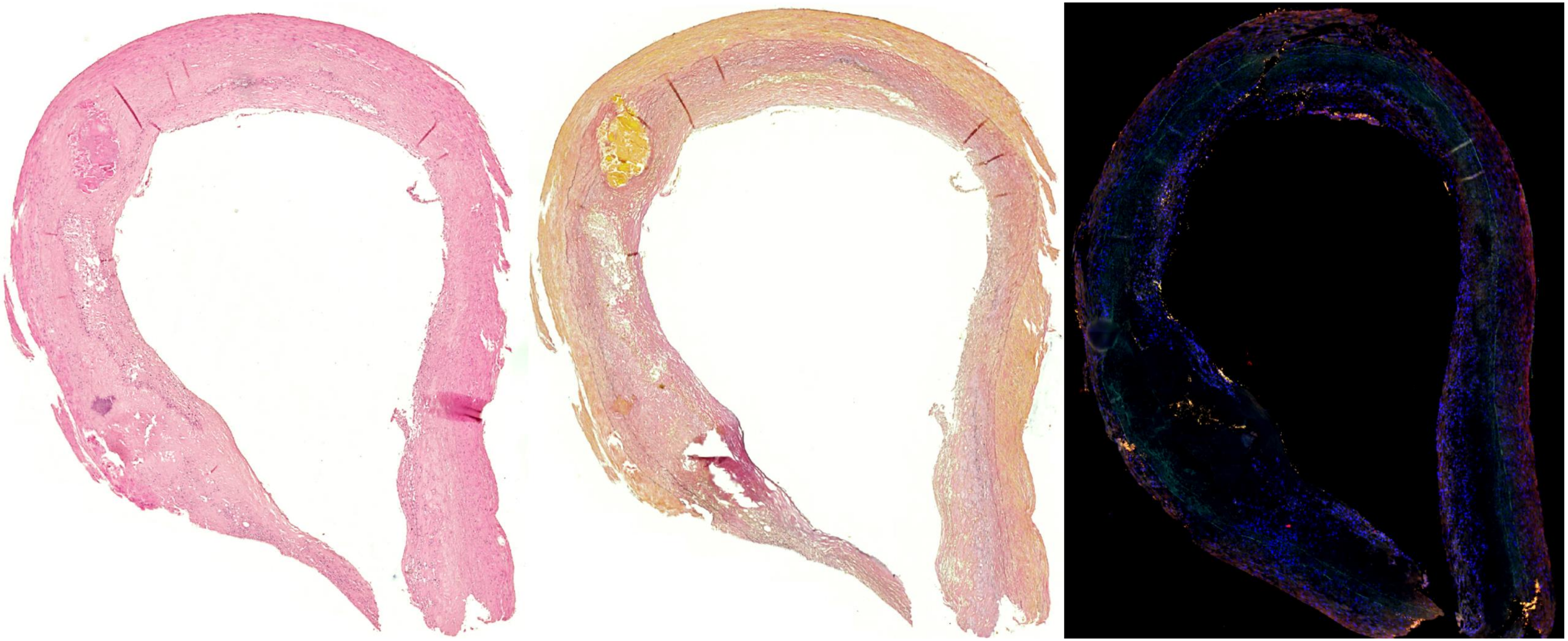

**Supplemental Figure 3** – Overview plaque of interest – different stainings. Coronary arteries and carotids in MISSION are processed in a standardized fashion using different staining methods to enable optimal histological characterization. As an example, an overview of a proximal LAD affected by atherosclerosis is shown here of a CHIP mutation carrier. The three images have been prepared from three immediate follow-up sections. **Left panel** - staining with HE. **Middle panel** - staining with EvG. **Right panel** - mutaFISH$^{TM}$ to screen for specific CHIP mutations.

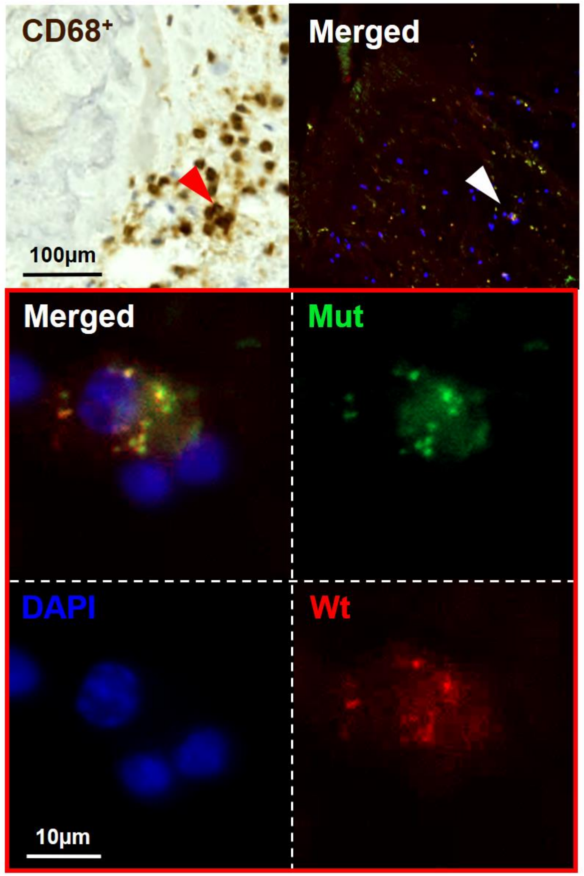


**Supplemental Figure 4** – Visualization of a single CHIP affected macrophage in the shoulder region of a human atherosclerotic plaque. **Upper panel** – the area of interest in this atherosclerotic plaque is stained for $CD68^{+}$ macrophages. Using mutaFISH™, staining for the specific DNMT3A mutation (Mut) c.2333T>G and wild type (Wt) at the RNA level was performed *in situ* in this atherosclerotic plaque of human FFPE tissue. **Lower panel** – provided are the merged and individual signals on single cell resolution. The DNMTA3 mutation c.2333G>T was detected via the green signal (Mut), the DNMT3A wild type via the red (Wt) signal and the cell nuclei (DAPI) via the blue signal. DAPI: 4′,6-Diamidin-2-phenylindole. mutaFISH: mutation-specific Fluorescence In Situ Hybridization.

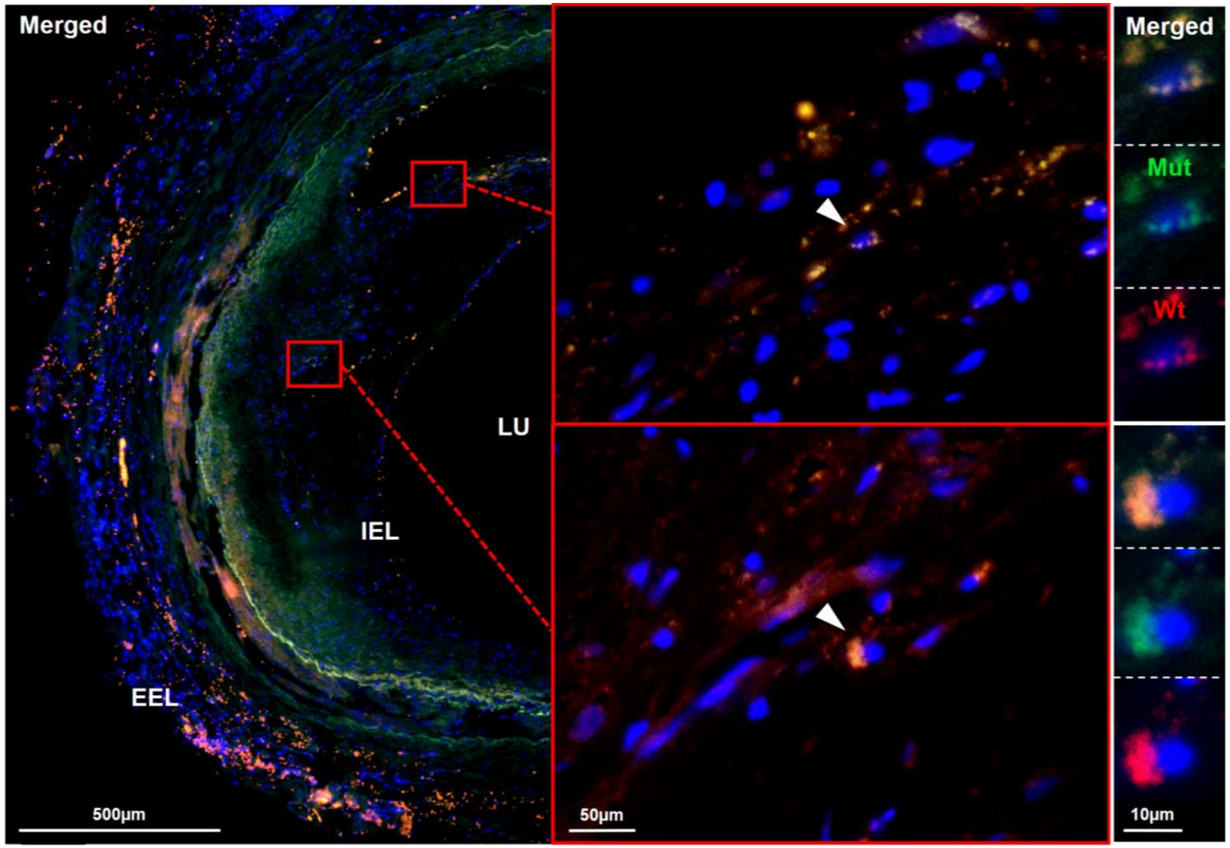


**Supplemental Figure 5** – DNMT3A CHIP mutation (c.2245C>T) in human atherosclerotic plaques. Staining for the specific DNMT3A mutation (Mut) c.2245C>T and wild type (Wt) at the RNA level was performed *in situ* in this advance atherosclerotic plaque of human FFPE tissue. **Left panel** – merged overview of the left part of the atherosclerotic plaque. Red boxes highlight the areas of interest. **Middle panel** – CHIP carrying macrophages are highlighted with white arrows. **Right panel** – provided are the merged and individual signals on single cell resolution. The DNMTA3 mutation c.2245C>T was detected via the green signal (Mut), the DNMT3A wild type via the red (Wt) signal and the cell nuclei (DAPI) via the blue signal. DAPI: 4′,6-Diamidin-2-phenylindole; EEL: external elastic lamina; FFPE: formalin-fixed paraffin embedded; IEL: internal elastic lamina; LU: lumen; mutaFISH: mutation-specific Fluorescence In Situ Hybridization.

### Adapted mutaFISH$^{TM}$ protocol

**Used Kits**

- mutaFISH RNA Probes KIT (Abnova Corporation, Taiwan)
- mutaFISH$^{TM}$ RNA Accessory KIT (KA4915, Abnova Corporation, Taiwan)

**Preparation of RNAse free buffers**

All buffers for the mutaFISH protocol have to be nuclease free.

- **Poly-L-Lysine 1:10:** Prepare 1:10 Poly-L-Lysine in ddH2O.
- **DEPC H2O:** Use 1 ml DEPC for 1000 ml ddH2O incubate for 1h at RT and autoclave.
- **DEPC PBS*:** Use 1 ml DEPC for 1000 ml PBS pH 7.4 incubate for 1h at RT and autoclave.
- **PBST*:** Use autoclaved DEPC PBS and add 1 ml Tween20 after autoclaving. If you use ready-to-use PBST Use DEPC and filter after 1h of incubation at RT.
- **Permeable Protease buffer:** Use 3mg/ml Pepsin to 0.5 M HCL.
- **Nuclease Free 1x Citric acid buffer pH6:** dilute nuclease free 10x ready-to-use Citric acid buffer in DEPC H2O or prepare 1x nuclease free buffer pH6.
- **2x SSC Buffer**:** Dilute ready-to-use nuclease free 20x SSC buffer in DEPC H2O or prepare 2x nuclease free buffer.
- **70% and 85% EtOH:** Dilute EtOH absolute to 70% and 85%.
- **3-4% Paraformaldehyde:** Paraformaldehyde solution has to prepared methanol free. Dilute in DEPC-PBS

* here Roti®fair PBST 7.4 and Roti® PBS 7.4 (CarlRoth GmbH&CoKG, Karlsruhe) were used

** here here Roti®-Stock 20x SSC (CarlRoth GmbH&CoKG, Karlsruhe) was used

**Protocol – Coating with Poly-L-Lysine**

1. Let Poly-L-Lysine (1:10 in ddH2O) come to room temperature.
2. Incubate Slides 7min at RT in **1:10 Poly-L-Lysine** for coating.
3. Remove slides from the rack and tap off water droplets.
4. Incubate slides at 56 °C for at least 1h.

**Protocol – mutaFISH**

Tissue preparation

1. Prepare 3-5 µm thick FFPE sections, air dry sections at heating plate (40 °C).
2. Incubate FFPE sections for 1h at 56 °C.

Deparaffinization and rehydration

1. Rinse slides 2 times in xylene substitute for each 5 min.
2. Immerse slides 2 times in 100% EtOH for each 3 min.
3. Immerse slides 2 times in 85% EtOH for each 3 min.
4. Immerse slides 2 times in 70% EtOH for each 3 min.
5. Wash slides in DEPC-H2O for 1 min and dry shortly at RT.

Target retrieval

1. Preheat heating plate to 75 °C.
2. Create secure bond with wax pen around the tissue sections.
3. Wash slides in DEPC-PBS for 2 min
4. Incubate slides with 1x citric buffer at 75-85 °C on heating plate for 20 min.
   *CAVE: Renew citric buffer every 10-15 min – sample should not dry out.*

5. Wash twice with 2x SSC buffer for 5 min.

*CAVE:Iff costaining with an antibody should be done perform permeabilization and immunostaining prior to mutaFISH and end up with 20 min fixation in 4% formaldehyde at RT.*

Fixiation and permeabilization

1. Incubate in 3-4% paraformaldehyde (provided) for 20 min at RT.
2. Immerse slides in 2x SSC buffer for 5 min.
3. Prewarm permeable buffer (provided in the kit or self-made) to 37 °C.
4. Use permeable buffer or 3 mg/mL Pepsin to 0.1 M HCl) at 37 °C for 30 min.
   *TIPP: RNAscope®* ***** *Protease III & Protease IV[3] reagents can be used. Here no prewarming is necessary.*
5. Wash slides in 2x SSC buffer for 5 min.

***** RNAscope® Protease IV (Advanced Cell Diagnostics, Inc., Canada) was used in this case

Dehydration

1. Immerse slides in 70% EtOH for 1 min.
2. Immerse slides in 85% EtOH for 1 min.
3. Immerse slides in 100% EtOH for 1 min.
4. Immerse slides in fresh PBST for 1 min.

*In situ* reverse transcription

1. prepare the following mixture on ice and use 100 µl of the mixture per slide.

| **Component** | **Amount per slide [µL]** |
|---|---|
| DEPC-H2O (Kit) | 60.5 |
| 5x RT Buffer (Kit) | 20.0 |
| BSA (Kit) | 1.0 |
| dNTP Mix (Kit) | 5.0 |
| **RT Primer (individual)** | 1.0 |
| RNase Inhibitor (Kit) | 2.5 |
| RT Enzyme (Kit) | 10.0 |
| **Total volume:** | **100.0** |

2. Incubate Slides at 37 °C in humidity oven over night.
   *CAVE: Take care that there is enough humidity. Slides should not dry out.*
3. Wash shortly in PBST.
4. Immerse slides with fresh PBST 2 times for each 2 min.

Postfixiation and probe hybridization

1. Cover tissue with 3-4% paraformaldehyde (provided) and incubate at 37 °C in humidity Box for 45 min.
   *CAVE: Check formaldehyde every 10-15 min – sample should not dry out.*
2. Wash slides with fresh PBST 2 times for 2 min.
3. prepare the following mixture on ice and use 100µl per slide.

| **Component** | **Amount per slide [µL]** | **Amount for negative control [µl]** |
|---|---|---|
| DEPC-H2O (Kit) | 32.5 | 34.5 |
| Formamide (Kit) | 20.0 | 20.0 |
| 10x Hybrid Enzyme Buffer (Kit) | 10.0 | 10.0 |
| 1 M KCl (Kit) | 5.0 | 5.0 |

| | | |
|---|---|---|
| **mutaFISH probe wt (individual)** | 1.0 | |
| **mutaFISH probe mutation (individual)** | 1.0 | |
| RNase Inhibitor (Kit) | 2.5 | 2.5 |
| RNaseH (Kit) | 8.0 | 8.0 |
| Hybrid Enzyme (Kit) | 20.0 | 20.0 |
| **Total volume:** | **100.0** | **200.0** |

4. Incubate at 37 °C in humidity oven for 60 min.
5. Heat up to 45 °C and incubate slides for another 90 min.
6. Immerse slide with fresh PBST 2 times for 2 min.

Amplification

1. Prepare the following mixture on ice and use 100 µl per slide.

| **Component** | **Amount per slide [µL]** |
|---|---|
| DEPC-H2O (Kit) | 61.5 |
| 50% Glycerol (Kit) | 10.0 |
| 10x DNA Polymerase Buffer (Kit) | 10.0 |
| BSA (Kit) | 1.0 |
| dNTP mix (Kit) | 5.0 |
| RNase Inhibitor (Kit) | 2.5 |
| DNA Polymerase (Kit) | 10.0 |
| **Total volume:** | **100.0** |

2. Incubate slides at 37 °C in humidity oven for 120 min.
3. Wash shortly in PBST.
4. Immerse slide with fresh PBST two times for 1 min.

Detection and counterstain

1. Use 100 µl of the following mixture per slide (prepare on ice).

| **Component** | **Amount per slide [µL]** |
|---|---|
| Detection Buffer (Kit) | 98.0 |
| **Detection probe for wt (individual)** | 1.0 |
| **Detection probe for mutation (individual)** | 1.0 |
| **Total volume:** | **100.0** |

2. Incubate at 37 °C in humidity oven for 60 min.
3. Wash shortly in PBST.
4. Immerse slide with fresh PBST 2 times for 2 min.
5. Immerse the slide in 70% EtOH for 0.5 min.
6. Immerse the slide in 85% EtOH for 0.5 min.
7. Immerse the slide in 100% EtOH for 0.5 min.
8. Mix 4 µl DAPI with 664 µl DEPC-PBS and apply 100 µl to the sample for 2-3 min at RT.

Sealing

- Immerse slide two times in fresh DEPC-PBS for 1 min.
- Cover slide with Prolong-Gold-Anti-Fade let it dry for 15 min and seal with nail-polish.
- Let dry slides for 1h and proceed with microscopy.
